# Published benefits of ivermectin use in Itajaí, Brazil for COVID-19 infection, hospitalisation, and mortality are entirely explained by statistical artefacts

**DOI:** 10.1101/2023.08.10.23293924

**Authors:** Robin Mills, Ana Carolina Peçanha Antonio, Greg Tucker-Kellogg

**Affiliations:** AI Horizon, The Netherlands; Adult Intensive Care Unit, Hospital de Clínicas de Porto Alegre, Brazil; Department of Biological Sciences, National University of Singapore, Singapore

## Abstract

**Background:** Two recent publications by Kerr et al. (Cureus 14(1):e21272; Cureus 14(8): e28624) reported dramatic effects of prophylactic ivermectin use for both prevention of COVID-19 and reduction of COVID-19-related hospitalisation and mortality, including a dose-dependent effect of ivermectin prophylaxis. These papers have gained an unusually large public influence: they were incorporated into debates around COVID-19 policies and may have contributed to decreased trust in vaccine efficacy and public health authorities more broadly. Both studies were based on retrospective observational analysis of city-wide registry data from the city of Itajaí, Brazil from July-December 2020.

**Methods:** Starting with initially identified sources of error, we conducted a revised statistical analysis of available data, including data made available with the original papers and public data from the Brazil Ministry of Health. We identified additional uncorrected sources of bias and errors from the original analysis, including incorrect subject exclusion and missing subjects, analysis of longitudinal data with cross-sectional design, an enrolment time bias, and multiple sources of immortal time bias. In models assuming no actual effect from ivermectin use, we conducted Monte Carlo simulations to estimate the contribution of these biases to any observed effect.

**Results:** Untreated statistical artefacts and methodological errors alone lead to dramatic apparent risk reduction associated with ivermectin use in both studies. The magnitude of apparent risk reduction from these artefacts is comparable to the results reported by the studies themselves, including apparent protection from infection, hospitalisation, and death, and including the reported apparent dose-response relationship.

**Conclusions:** The inference of ivermectin effect reported in both papers is unsupported, as the observed effects are entirely explained by untreated statistical artefacts and methodological errors. Our re-analysis calls for caution in interpreting highly publicised observational studies and highlights the importance of common sources of bias in clinical research.

## Introduction

The first half of 2020 was marked by both the beginning of the COVID-19 pandemic and frantic efforts around the globe to prevent and contain its spread. Most of those initial efforts relied on non-pharmaceutical interventions (e.g., social distancing, masks, travel restrictions, and regional lockdowns) to “flatten the curve” and reduce the strain on healthcare systems before effective treatments and vaccines became available [1, 2]. As the pandemic unfolded, healthcare systems worldwide rapidly became overburdened with an increasing number of severely ill patients and high COVID-19 mortality rates.

In Brazil, there was immense interest in early COVID-19 treatment during the initial phase of the pandemic, including the potential use of the anti-parasitic drug ivermectin [3]. While there was no clinical evidence of ivermectin efficacy for COVID-19, initial *in vitro* studies at the time had shown potential antiviral activity of ivermectin in cell culture [4], which fuelled interest in its use. Starting in July 2020, the city of Itajaí (in the southern Brazil state of Santa Catarina) [5, 6] implemented a controversial city-wide program in July 2020 as a potential COVID-19 prophylaxis. Eligible residents were offered ivermectin pills with an intermittent dosing schedule of 0.2 mg/kg of body weight (up to a maximum 24 mg for those above 90 kg body weight) each day for two consecutive days, repeated every 15 days.

In two closely related retrospective analysis studies of the Itajaí program, Kerr, Cadegiani et al. [7, hereafter KC22] and Kerr, Baldi et al. [8, hereafter KB22] made dramatic claims of ivermectin benefit. KC22 concluded that using ivermectin in the Itajaí program resulted in a 44% reduction in COVID-19 infections. Among all infected individuals, KC22 reported 37% reduction in hospitalisation and 43% reduction in mortality associated with ivermectin use. These already dramatic results were even larger after the application of propensity score matching (PSM) and adjustment for other covariates. Furthermore, KB22 presented an even more startling dosedependent benefit among infected individuals: the so-called “strictly regular” ivermectin users experienced a 92% reduction in mortality compared to non-users and an 82% reduction compared to irregular users.

These two papers gained significant public attention and contributed to ongoing public policy debates, both about COVID-19 treatment and prevention, and about trust in the medical and scientific establishment. Each paper has an Altmetric score in the top five per cent of *all* scientific research, has been the subject of news reporting, and has been shared on social media or viewed at the journal site hundreds of thousands of times. KC22 and KB22 have also attracted the attention of fact-checkers, but published critiques to date have mostly focused either on the limits of observational studies in general or on missing covariates and superficial peer review of these papers specifically [9–11]. In response, the Editor in Chief of Cureus has defended the peer review process and subsequent publication of these studies [12].

Given the widespread dissemination and discussion of these papers, it is crucial to provide an unbiased critique to foster a more informed public debate on scientific and medical research, ultimately helping to protect the public from scientific misinformation [13, 14]. This unbiased critique is particularly significant for influential papers addressing polarising topics of public interest, such as the use of ivermectin as a treatment for COVID-19 [15].

In this work, we take a direct approach by reanalysing the data available from KC22 and KB22 and combining it with public data from the Brazilian Health Ministry. We identify a variety of important statistical fallacies and other errors, and use simulations to estimate the consequences of leaving these issues untreated. Our analysis demonstrates that the seemingly dramatic benefits of prophylactic ivermectin for COVID-19, as reported in both KC22 and KB22, can be entirely attributed to unresolved statistical fallacies present in the original analyses. The code for all analyses presented in this manuscript can be found on GitHub, ensuring the reproducibility of our findings.

## Results

### The data from KC22 and KB22

The analyses of KC22 and KB22 compared events (infections, hospitalisations, and deaths) between participants in the program (who volunteered to take ivermectin as a *prophylactic* agent) and non-participants within a fixed study period (from July 7 through December 2, 2020). KC22 described the data set as *excluding* individuals who tested positive from the registry data prior to July 7, 2020.

KC22 combined two data sets: one of the participants in the ivermectin prophylaxis program and the other from a citywide population registry to retrieve non-participants’ data. As we received no response about the availability of original data sets after contacting both the city authorities in Itajaí and the authors of KC22 and KB22, we restricted our analysis to KC22 supplementary data on OSF posted with KC22 by the corresponding author Flavio Cadegiani and official public data from the Brazil Ministry of Health.

KC22 claimed virtually no missing values in the data because of mandatory reporting and indeed, most fields for each individual record were completely filled in. However, crucial information was missing for *all* participants in the data. For example, the amount of medicine for an individual was a function of weight (≤0.2 mg ivermectin/kg body weight/day for two days every 15 days), but neither body weight nor dosage category were reported. The KC22 data also failed to include any dates other than the date of birth; dates of program enrolment, medication collection, infection, hospitalisation, or death were all missing.

Data entry errors in KC22 data were not uncommon. For example, while the maximum possible total ivermectin usage over the study period was 80 tablets, hundreds of users were recorded as having more than 80, and in some cases thousands, of tablets.

### KC22 mistakenly included prior infections and hospitalisations, primarily in the non-user group

The authors describe the data as representing “all events” of COVID-19 infection, hospitalisation (in public hospitals), and deaths within the study period (7 July and 2 December 2020). The dates of these events, however, were neither used in the original analysis nor contained in the supplementary data. Fortunately, official public data from the Brazilian Health Ministry’s Unified Health System (DATASUS) provides detailed information for hospitalisations and deaths of COVID-19 patients across Brazil, including event dates. To understand the distribution of events in the KC22 data, we matched city-level Itajaí data with records from the KC22 data based on the date of birth, sex, and 2020 COVID-19 hospitalisation and death.

The majority of recorded deaths and hospitalisations in KC22 were matched to DATASUS data (132/141 deaths, 94%, and 43/93 hospitalised survivors, 46%). To our surprise, COVID-19 symptom onset occurred before July 7, 2020 in 45 of the matched individuals, and 87% of those (39/45) were classified as non-users in the KC22 analysis (Table 1A, p < 0.001 for association between exposure group and mistaken inclusion). When we looked at subgroups by mortality for those individuals, the bias for mistaken inclusion of early infections in the non-user group was more severe for those who died (31 non-users, 2 users, 94% non-user, p < 0.001) than for those who survived (8 non-users, 3 users, 73% non-user, p = 0.088Table 1B).

**Table 1.** Mistaken inclusion of pre-study infections in the non-user group. KC22 study participants uniquely mapped with Brazil Health Ministry (SUS) individuals as described in the text. Parts (a) and (b) show overall and subgroup analysis, respectively.

|  | non-user<sup>1</sup> | IVM user<sup>1</sup> | p-value<sup>2</sup> |
| --- | --- | --- | --- |
| **All Brazilian Health Ministry-mapped individuals (178 total)** |  |  | **<0.001** |
| Covid onset on or after 7 July 2020 | 58 (60%) | 72 (92%) |  |
| Covid onset before 7 July 2020 | 39 (40%) | 6 (7.7%) |  |

<sup>1</sup> n (%)
<sup>2</sup> Fisher's exact test

|  | non-user<sup>1</sup> | IVM user<sup>1</sup> | p-value<sup>2</sup> |
| --- | --- | --- | --- |
| **Alive (43)** |  |  | **0.088** |
| Covid onset on or after 7 July 2020 | 13 (62%) | 19 (86%) |  |
| Covid onset before 7 July 2020 | 8 (38%) | 3 (14%) |  |
| **Dead (135)** |  |  | **<0.001** |
| Covid onset on or after 7 July 2020 | 46 (60%) | 56 (97%) |  |
| Covid onset before 7 July 2020 | 31 (40%) | 2 (3.4%) |  |

<sup>1</sup> n (%)
<sup>2</sup> Fisher's exact test

Because the DATASUS records during the study period are primarily hospitalisations and deaths, we do not currently have strong direct evidence that the biased inclusion of pre-study infections in the non-participant group extends to non-hospitalised subjects. However, the mistaken pre-study enrolment of non-users accounted for 40% of matched non-users, and all infected non-users were included in the propensity score matching of KC22, so the impact of this mistake alone was dramatic.

### KC22 data was biased towards a subset of early infections

KC22 and KB22 under-reported deaths and severely under-reported hospitalisations from COVID-19 in Itajaí. While KC22 reports 141 deaths, DATASUS records 186 adult COVID-19 deaths among Itajaí residents during the study period, and 183 reported deaths following symptom onset in the same period, 30% higher than in KC22. KC22 reported 185 total hospitalisations from a study population of 159,560 (0.12% hospitalised); official government data reports 651 Itajaí resident adult hospitalisations for COVID-19 from infections in the second half of 2020, of whom 479 had symptom onset during the KC22 study period (a hospitalisation rate 259% greater than that reported in KC22).

The death/hospitalisation ratio was also much higher for KC22 (141/185, 76.1%) than for Itajaí as a whole (183/479, 38.5%). KC22 included only hospitalisations in public hospitals, which may partially account for the difference among hospitalised survivors and death/hospitalisation ratio but raises fundamental unaddressed issues of confounding. The under-reporting of hospitalisations and deaths cannot be attributed to the reported ivermectin effects claimed by KC22.

To further understand this issue, we compared the dates of COVID-19 symptom onset, hospitalisation, and death for the 175 records mapped between KC22 and DATASUS (Fig. 1A, top). Dates of death were almost entirely contained within the study period, consistent with KC22’s claim to have analysed events during this period. The same is true of hospitalisation dates for hospitalised survivors. However, dates of hospitalisation for matched deaths (Fig. 1A, middle) and dates of symptom onset for both hospitalisations and deaths (Fig. 1A, bottom) occurred well before the beginning of the study period.

**Figure 1.**
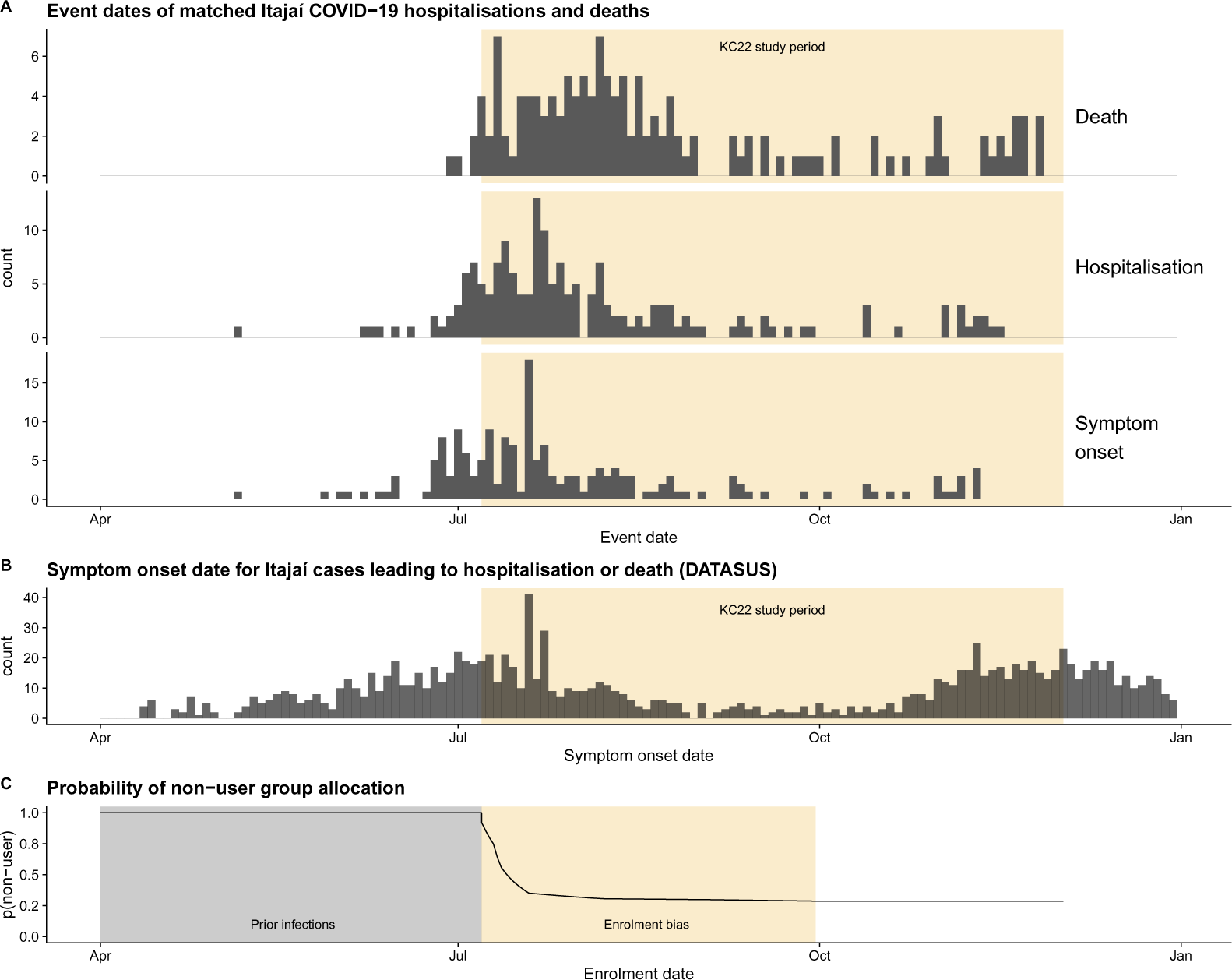
Symptom onset and study group allocation. A. Event dates for 132 matched Covid deaths and 43 matched hospitalised survivors mapped to the SUS data from KC22. The three sub-panels show recorded dates of death, hospitalisation, and symptom onset. B. Official Brazilian Health Ministry entries for records of death or hospitalisation among adult residents of Itajaí. C. The probability of an individual considered in the study being allocated to the non-user group changes dramatically over time during the allocation of participants in KC22. Enrolment was estimated using data from contemporaneous Itajaí municipal bulletins and news reporting. See Methods for details.

Strikingly, the KC22 data not only mistakenly included hospitalisations and deaths with symptom onset *before* the study period, but almost all of the remaining matched hospitalised individuals experienced symptom onset in the first half of the study period (Fig. 1A, bottom). KC22 thus almost entirely neglected the second wave of COVID hospitalisations, which peaked at the end of the study period (Fig. 1B). Indeed, symptom onset among the *uncounted* deaths primarily occurred at the tail end of the study period, when the December 2020 wave was getting under way (Fig. S1).

The KC22 analysis appears to treat the data as cross-sectional, analysing (as described in KC22) “all events as they occurred during the citywide governmental COVID-19 prevention with ivermectin program, from July 7, 2020, to December 2, 2020”. In reality, the data is inherently longitudinal: hospitalisations precede deaths and infections precede hospitalisations. Including all deaths within a fixed time period, as described in KC22, would lead to mistaken inclusion of prior hospitalisations and infections (as seen in Fig. 1A); including all hospitalisations would have inevitably included prior infections. The opposite problem would appear near the end of the study period, where hospitalisations and deaths would be excluded from analysis even if the infections that preceded them occurred during the study period. This is exactly what is observed (see Fig. S1), in addition to the under-counting of events late within the study period.

### Enrolment to the ivermectin program continued during the study period

Both KC22 and KB22 compared events between adult residents of Itajaí who did or did not participate in the ivermectin program over the entire study period. KC22 claimed “[t]his strict interval avoids differences in terms of periods of exposure”. This *cannot* be true, however, as not all 113,845 participants started exactly on July 7, 2020. Consider an individual who reported symptoms on July 8, but had not yet joined the ivermectin distribution program. That individual would be ineligible for inclusion in the KC22 study as an ivermectin user, but would instead be treated in the analysis of KC22 as an infected non-user. The biased allocation of new infections into the non-user group (Fig. 1C) would continue for as long as the distribution program enrolled new participants. This alone is a classic case of immortal time bias [16], but because the most rapid enrolment occurred during the peak infection period in July 2020 (Fig. 1B), it also coincides with substantial chronological bias [17]. We describe the immortal time bias and chronological bias during enrolment collectively as “enrolment bias” to distinguish these two sources of bias from other sources of bias described later.

### Enrolment bias, incorrect inclusion of already infected participants, and biased sampling led to large apparent protection against infection, hospitalisation, and death in KC22

We estimated the effects of the issues above using Monte Carlo simulation. We simulated the KC22 study under a model of no actual ivermectin effect but successively including enrolment bias, the incorrect inclusion of subjects with symptomatic onset prior to the study, and the empirically biased sampling towards the beginning of the study. We simulated symptom onset in individual patient data cohorts each the size of the data set in KC22. To simulate the dates of symptom onset, we used daily Itajaí notifications of infection from the Brazil Health Ministry. To simulate enrolment over time, we used contemporaneous local municipal bulletins and news reports of program enrolment and assumed participants began taking ivermectin immediately.

We developed multiple models to estimate the individual and cumulative effects of the identified biases and errors. Each model used the numbers reported in KC22:7231 infections within a cohort of 159,560 individuals (infection rate of approximately 4.5%). Additionally, we simulated hospitalisations and deaths to match KC22 totals, using dates sampled from the Brazil Health Ministry records, as detailed in the Methods and Materials section. Results are reported based on 1000 simulations of each model and compared to reported values from KC22. Hospitalisations and deaths reported among infected individuals and compared to the infected individuals from KC22 prior to propensity score matching.

The enrolment bias present in KC22 leads to an estimated fictitious risk reduction for infection of 17% for users of an ineffective medicine. This result stems from our first model (i-ENR), in which we randomly sampled infections from the distribution of onset dates between July 7 and December 2, 2020. In addition to the enrolment bias, the incorrect inclusion of subjects with symptomatic onset prior to the study period leads to an additional 10% fictitious risk reduction for infection. In our second model (i-INF), we simulated incorrect inclusion by sampling from symptom onset dates where the notification date was between July 7 and December 2, 2020. The i-INF model thus encompassed both enrolment bias and the mistaken inclusion of early infections.

Lastly, an additional 24% fictitious risk reduction for infection is caused by the biased sampling towards the beginning of study observed in KC22. In the third model family (i-KC22), we simulated the sampling bias in Fig. 1B by sampling infections using the distribution of infection dates of matched individuals. Details are discussed in the methods section. The i-KC22 model family most closely matched the errors and biases so far discussed in KC22.

The results of the models, alongside the published results of KC22 and a trivial negative model (i-NEG, with neither ivermectin effect nor any biases) are shown in Table 2. Each source of bias reduced the apparent incident rates of infection, hospitalisation, and death of ivermectin users and, hence, increased the apparent risk reduction of the exposure. Biases in the design and execution of KC22 account for virtually all of the reported protection attributed to ivermectin.

**Table 2.** Apparent protection provided by biases and errors in KC22. Apparent protection against infection, hospitalisation, and death due to enrolment bias, mistaken inclusion of prior infections, and missing data. Comparisons are between users and non-users, as defined in KC22. Three different simulation strategies were used to simulate the isolated effects of biases identified in [7] along with a negative control (i-NEG). i-KC22 and the published KC22 data set include all three biases. Simulations are as described in the text.

|  | Simulation |  |  | Literature |  |
| --- | --- | --- | --- | --- | --- |
|  | i-NEG | i-ENR | i-INF | i-KC22 | KC22 [7] |
| has enrolment bias |  | ✓ | ✓ | ✓ | ✓ |
| includes prior infections |  |  | ✓ | ✓ | ✓ |
| has missing data |  |  |  | ✓ | ✓ |
| **Infection** |  |  |  |  |  |
| Rate (non-user) | 4.53% | 5.14% | 5.62% | 7.15% | 6.64% |
| Rate (user) | 4.53% | 4.29% | 4.10% | 3.48% | 3.69% |
| Risk ratio |  | 0.834 | 0.729 | 0.487 | 0.555 |
| 95% CI |  | [0.80–0.88] | [0.70–0.76] | [0.47–0.51] | [0.53–0.58] |
| *p* |  | <0.001 | <0.001 | <0.001 | <0.001 |
| Risk reduction |  | 17% | 27% | 51% | 44% |
| **Hospitalisation** |  |  |  |  |  |
| Rate (non-user) | 0.12% | 0.14% | 0.16% | 0.20% | 0.22% |
| Rate (user) | 0.12% | 0.11% | 0.09% | 0.08% | 0.08% |
| Risk ratio |  | 0.750 | 0.588 | 0.398 | 0.349 |
| 95% CI |  | [0.56–1.03] | [0.44–0.80] | [0.28–0.56] | [0.26–0.47] |
| *p* |  | 0.039 | <0.001 | <0.001 | <0.001 |
| Risk reduction |  | 25% | 41% | 60% | 65% |
| **Death** |  |  |  |  |  |
| Rate (non-user) | 0.09% | 0.11% | 0.13% | 0.17% | 0.17% |
| Rate (user) | 0.09% | 0.08% | 0.07% | 0.06% | 0.05% |
| Risk ratio |  | 0.707 | 0.541 | 0.335 | 0.315 |
| 95% CI |  | [0.50–1.02] | [0.39–0.77] | [0.25–0.47] | [0.22–0.43] |
| *p* |  | 0.026 | 0.002 | <0.001 | <0.001 |
| Risk reduction |  | 29% | 46% | 66% | 68% |

The estimated effect of enrolment bias in the i-ENR model is conservative, since we assumed optimistically that news reporting was accurate and that ivermectin use began immediately for all participants. In addition, we did not include uncertainty in the model parameters, so confidence intervals are likely under-estimated. While we have direct evidence for the temporal-biased sampling in the case of hospitalisation and death, the effect of temporal sampling bias for infections in i-KC22 is indirectly estimated based on assumed consistency with hospitalisations and deaths. Full details of the simulation methods are found in the Methods and in the GitHub repository accompanying this paper.

### KC22 hospitalisation and mortality results introduced attrition bias by design

COVID-19 outcome events occur in sequence: hospitalisation usually precedes COVID-19 death; COVID-19 symptom onset always precedes COVID-19 hospitalisation and COVID-19 death. KC22 stated they had included “all events” from July 7–December 2, 2020, so hospitalisations and deaths *after* the study period following symptom onset *during* the study period were ignored. The enrolment bias described above thus leads not just to a fictitious protection against infection, but to differences between exposure groups in the distribution of infections over time: non-user infections accumulate earlier in the study period, and infections among users accumulate later. Indeed, the median symptom onset date for the non-user group over 1000 simulations of i-ENR was September 4, 2020, while that for the user group was October 2, almost a month later. The temporal difference in symptom onset leads to additional bias for later hospitalisations and deaths due to attrition [18, 19].

Attrition from events after the study period can occur in both exposure groups. We estimated attrition rate from failure to count post-study events in each exposure group in all simulation models, and calculated a risk ratio for hospitalisation and death within each group by comparing the case without attrition (no cutoff for event counting) to the case with attrition (events after December 2, 2020 not counted). As shown in Table 3, attrition bias provides apparent (though fictitious) protection against hospitalisation and death in all cohorts, and in all cases the fictitious protection is greater in the user group than the non-user group. Adding the possibility of pre-study inclusion in the i-INF model (Table 3B) lowers the protection in the non-user group when compared to the enrolment-bias only i-ENR model. Attrition bias in the i-KC22 model was estimated by including unmatched late deaths (Fig. S1) in the model.

**Table 3.**
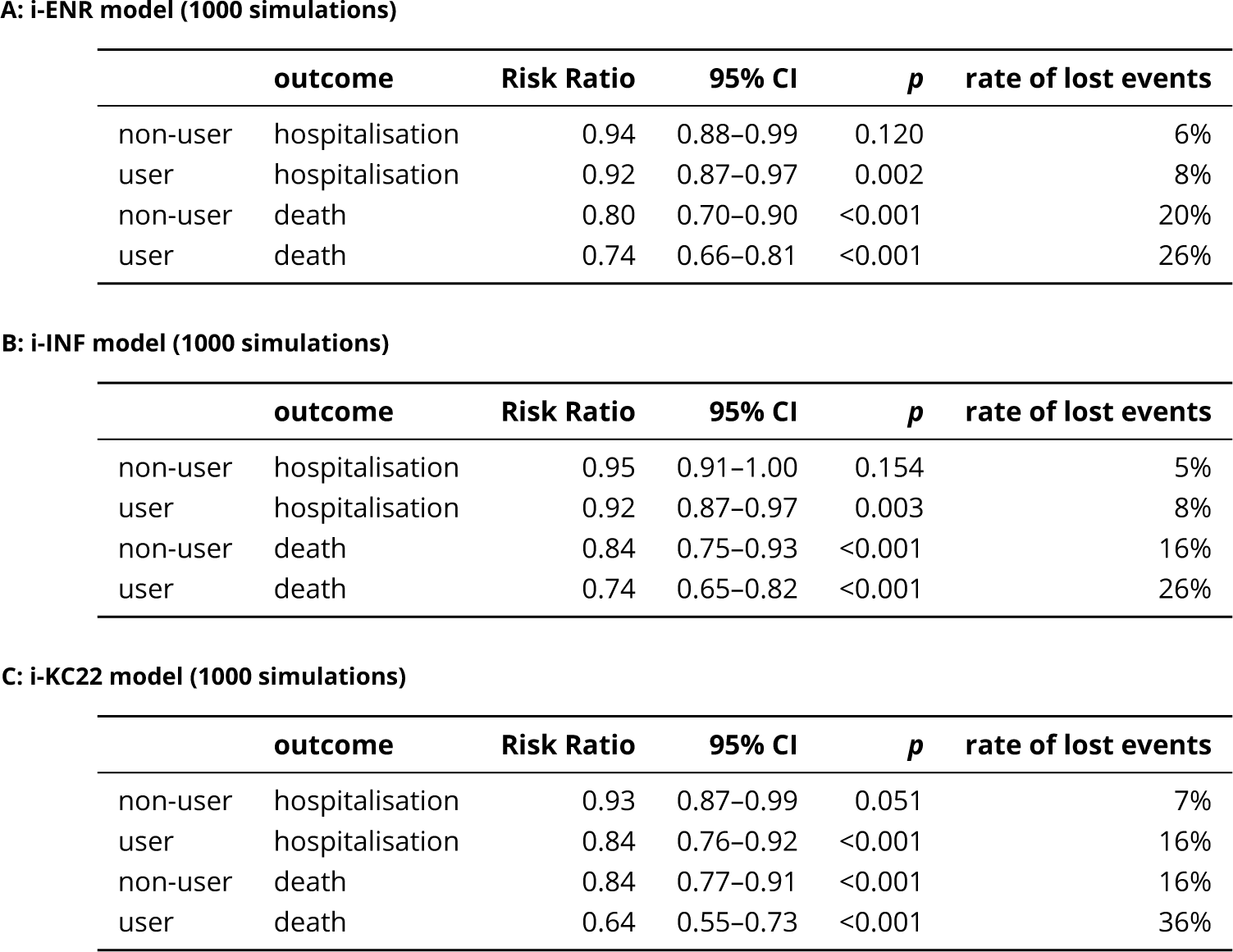
Attrition bias provides apparent protection against hospitalisation and death in all cohort. Risk ratios indicate apparent protection from adverse outcomes by loss of events occurring after the study period. Note that the apparent protection in all models is greater for death than for hospitalisation, and greater in the user group than in the non-user group.

**A: i-ENR model (1000 simulations)**
|  | outcome | Risk Ratio | 95% CI | p | rate of lost events |
| --- | --- | --- | --- | --- | --- |
| non-user | hospitalisation | 0.94 | 0.88–0.99 | 0.120 | 6% |
| user | hospitalisation | 0.92 | 0.87–0.97 | 0.002 | 8% |
| non-user | death | 0.80 | 0.70–0.90 | <0.001 | 20% |
| user | death | 0.74 | 0.66–0.81 | <0.001 | 26% |

**B: i-INF model (1000 simulations)**
|  | outcome | Risk Ratio | 95% CI | p | rate of lost events |
| --- | --- | --- | --- | --- | --- |
| non-user | hospitalisation | 0.95 | 0.91–1.00 | 0.154 | 5% |
| user | hospitalisation | 0.92 | 0.87–0.97 | 0.003 | 8% |
| non-user | death | 0.84 | 0.75–0.93 | <0.001 | 16% |
| user | death | 0.74 | 0.65–0.82 | <0.001 | 26% |

**Table 3. Attrition bias provides apparent protection against hospitalisation and death in all cohort.**
|  | outcome | Risk Ratio | 95% CI | p | rate of lost events |
| --- | --- | --- | --- | --- | --- |
| non-user | hospitalisation | 0.93 | 0.87–0.99 | 0.051 | 7% |
| user | hospitalisation | 0.84 | 0.76–0.92 | <0.001 | 16% |
| non-user | death | 0.84 | 0.77–0.91 | <0.001 | 16% |
| user | death | 0.64 | 0.55–0.73 | <0.001 | 36% |

Fig. 2 illustrates the basis of this finding in detail. In KC22, infections tended to occur earlier in the non-user group because of the delayed enrolment in the user group and the July 2020 infection peak (Fig. 2A). Death events after the follow-up period are lost to attrition as a rule in any given cohort, but this attrition is more likely among users than non-users in KC22, as shown for a single simulated cohort in Fig. 2B. A higher percentage of deaths than hospitalisations are lost to attrition in both groups (Fig. 2C) but the ivermectin user group is more likely to have uncounted hospitalisations and deaths than the non-user group. This translates to an increased risk of attrition in the ivermectin group, a relative risk that is higher for deaths than for hospitalisations (Fig. 2D) Note that the protection shown in Fig. 2D is relative protection of users compared to non-users, as opposed to the apparent protection provided from attrition within each group shown in Table 3. Attrition bias for the i-INF and i-KC22 models are shown in Figs. S2 and S3, respectively.

**Figure 2.**
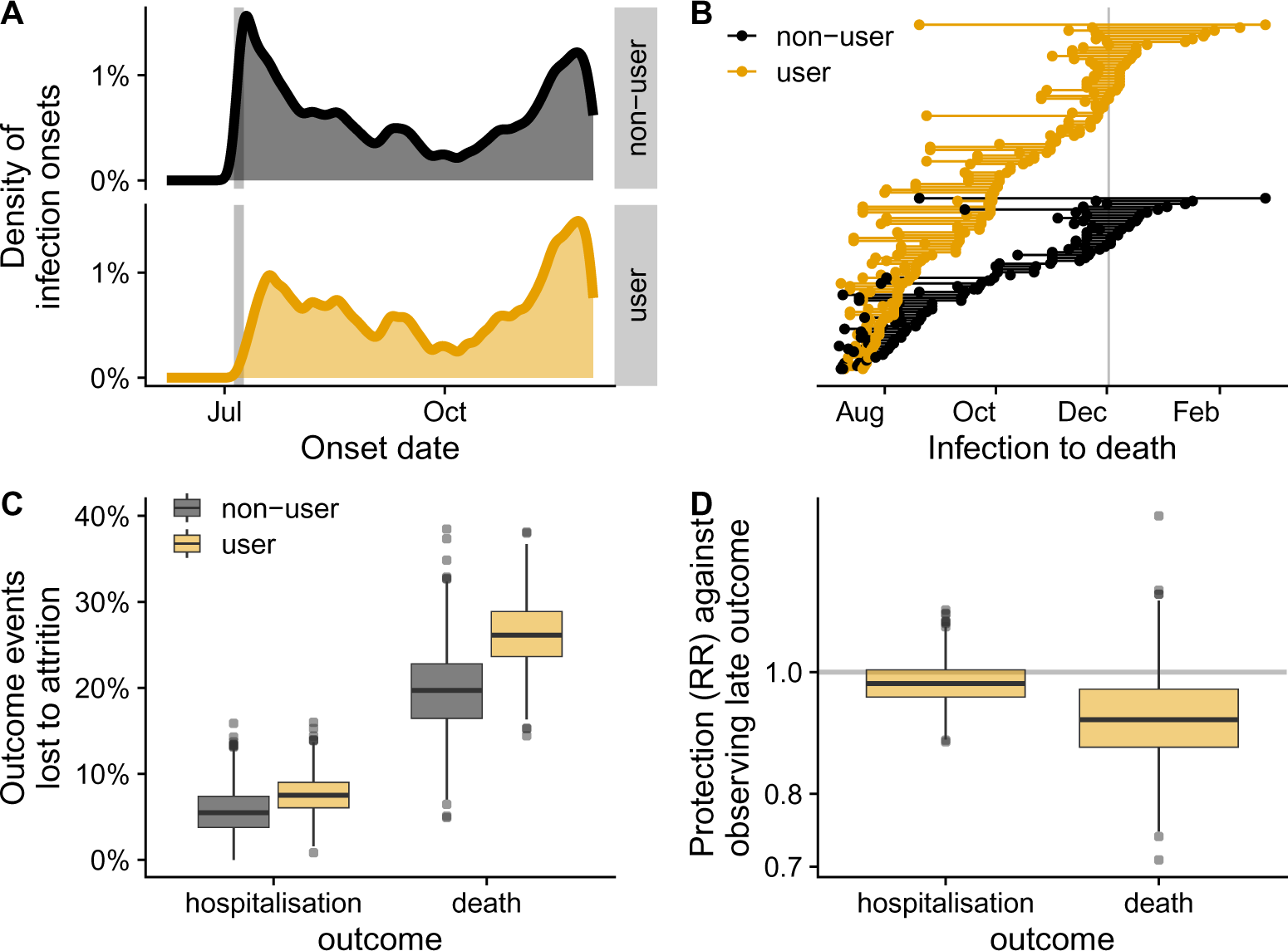
Attrition bias in hospitalisations and deaths in KC22 stemmed from enrolment bias. Results are shown for the iENR simulation model; A. Empirical distributions of simulated infection dates over 1000 runs of the i-ENR model. Note the delayed early peak of infections in the ivermectin user group. B. Example from one typical simulation of uncounted deaths among ivermectin users. Each line segment represents an individual in the simulation who was infected and later died, with infection and death dates at the end points. The study end date is marked with a vertical line. C. Hospitalisations and deaths are lost to attrition more frequently in the user group (1000 simulations of the i-ENR model). D. The ivermectin user group has apparent (but fictitious) protection from hospitalisation and death compared to non-users because hospitalisations and deaths are more likely to occur after the end of the study period for the user group. (Results under the other models are shown in Figures S2-S3.)

### The “regular ivermectin user” distinction in KB22 created additional immortal time bias

We now turn to the second paper from the Itajaí study (KB22), which considered the “regular use” of ivermectin. Because actual ivermectin use was not measured, KB22 treated the return to collect medication over time as a surrogate measure for the actual ivermectin intake. KB22 further subdivided ivermectin users into exposure groups based on the total amount of medication distributed: regular users (those who had received at least thirty 6 mg ivermectin tablets, or 180 mg total), irregular users (those who had received no more than 10 tablets), and non-users. According to KC22, newly diagnosed COVID-19 patients were recommended “not to use ivermectin” and that “The city did not provide or support any specific pharmacological outpatient treatment for subjects infected with COVID-19”. The citywide ivermectin program was, as KC22 and KB22 made clear, the use of ivermectin as a prophylactic for COVID-19, not as a treatment.

The central claim of KB22 was a dose-response relationship between ivermectin use and protection from infection, hospitalisation, and death. The exposure groups in KB22, however, were assigned retrospectively based on the amount of medication distributed over time. Moreover, both the language of KC22 and contemporary news reports suggest that users would have stopped ivermectin use upon infection, but the analysis of KB22 assumes the opposite, treating ivermectin usage as an independent variable.

How long would someone have to take ivermectin to be classified as a regular user in KB22? The maximum dosage in the Itajaí program was 4 tablets a day for two days, or 8 tablets every 15 days from day 2 of usage. This would require a minimum of 46 days after enrolment to be classified as a regular user. The more typical 3-tablet dosage would require 61 days. This entire time period is “immortal” time for regular users who stopped ivermectin use upon infection: infections during that time period would result in their allocation to other groups and an apparent reduction of the infection rate in regular users.

We simulated the effects of these biases by randomly allocating intended usage among ivermectin users using the total tablet distribution from KC22 data, and truncating to simulated actual usage based on how long an individual participated in the study without infection. We assumed a body weight between 60-90 kg, so 3 tablets per day of use. Because actual changes in usage upon infection are unknown, we considered two distinct scenarios. In the deterministic scenario, all infected users stopped ivermectin upon infection. In the probabilistic scenario, we used the regularity groupings of KB22 as a proxy for commitment: regular users receiving ≥30 tablets would stop on infection with a probability of 5%, irregular users would stop with a probability of 30%. As there is no evidence that ivermectin was offered to COVID-19 inpatients at the time, all users would stop taking ivermectin on hospitalisation.

The immortal time bias provided by the “regular user” definition in KB22 leads to a strong fictitious dose-response relationship under the assumption of ineffective medicine (Figures 3, S4, and S5.) In the deterministic stopping scenario the dose-response relationship was even stronger than reported in KB22. In the probabilistic setting, which models a very small change in user behaviour upon infection, our simulations closely match the dose-response relationship found in KB22 (see Table 4).

**Figure 3.**
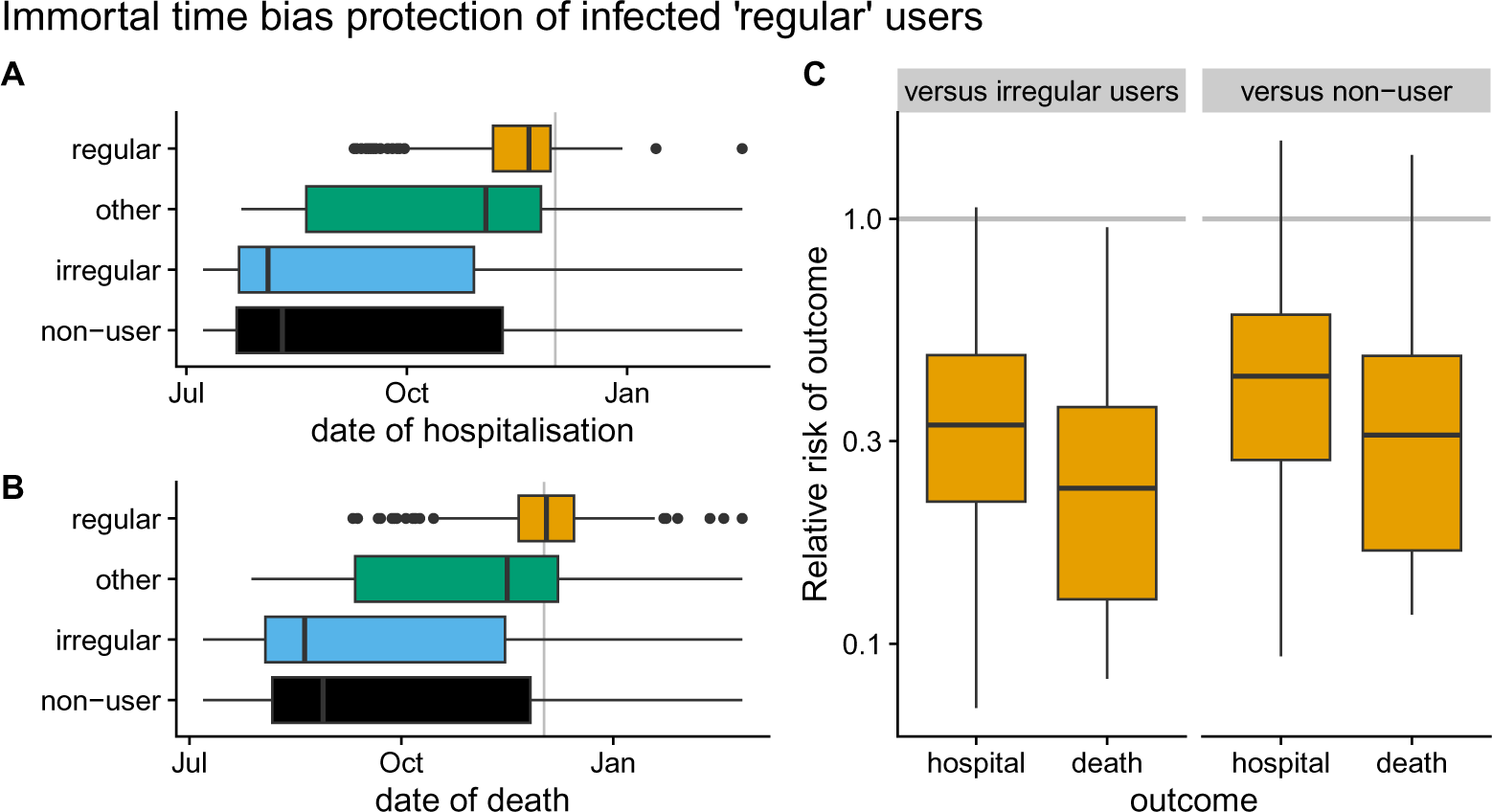
KB22’s “regular” use group [8] created more immortal time bias. A. Simulated dates of hospitalisation in i-ENR for individuals grouped by exposure according to KB22-defined usage groups. The study end date is marked with a vertical line. B. Dates of death. C. Relative risk of hospitalisation and death for “regular” ivermectin users compared to “irregular” and non-users. Risk ratios are calculated and plotted for each of 1000 simulations. Dates of hospitalisation and death are shown for all individuals across 1000 simulations. (Corresponding results under the i-INF and i-KC22 models are shown in Figures S4 and S5, respectively.)

**Table 4.** i-KC22 model with probabilistic stop on infection. Stop probability: 0.30 (irregular), 0.05 (regular). Statistics for hospitalisations and deaths are reported for all individuals in the cohort.

| Summary statistics | Simulation |  |  |  | KB22/KC22 |  |  |  | 'Corrected' |  |  |  |
| --- | --- | --- | --- | --- | --- | --- | --- | --- | --- | --- | --- | --- |
|  | RR | 95% CI | Risk Red. | p.val | RR | 95% CI | Risk Red. | p.val | RR | 95% CI | Risk Red. | p.val |
| **Infection** |  |  |  |  |  |  |  |  |  |  |  |  |
| non-user vs user | 0.49 | 0.47–0.51 | 51% | <0.001 | 0.56 | 0.53–0.58 | 44% | <0.001 | 1.14 | 1.07–1.22 | -14% | 1.000 |
| non-user vs irregular | 0.64 | 0.60–0.68 | 36% | <0.001 | 0.68 | 0.64–0.73 | 32% | <0.001 | 1.07 | 0.98–1.16 | -7% | 0.987 |
| non-user vs regular | 0.37 | 0.33–0.43 | 63% | <0.001 | 0.51 | 0.45–0.57 | 49% | <0.001 | 1.38 | 1.15–1.65 | -38% | 1.000 |
| irregular vs regular | 0.59 | 0.51–0.67 | 41% | <0.001 | 0.75 | 0.66–0.85 | 25% | <0.001 | 1.29 | 1.07–1.55 | -29% | 0.999 |
| **Hospitalisation** |  |  |  |  |  |  |  |  |  |  |  |  |
| non-user vs user | 0.40 | 0.28–0.56 | 60% | <0.001 | 0.35 | 0.26–0.47 | 65% | <0.001 | 0.93 | 0.58–1.41 | 7% | 0.294 |
| non-user vs irregular | 0.74 | 0.49–1.09 | 26% | 0.045 | 0.52 | 0.34–0.74 | 48% | <0.001 | 0.76 | 0.42–1.27 | 24% | 0.073 |
| non-user vs regular | 0.15 | 0.01–0.39 | 85% | <0.001 | 0.00 | 0.00–0.00 | 100% | <0.001 | NaN | NaN | NaN | NA |
| irregular vs regular | 0.21 | 0.01–0.56 | 79% | <0.001 | 0.00 | 0.00–0.00 | 100% | <0.001 | NaN | NaN | NaN | NA |
| **Death** |  |  |  |  |  |  |  |  |  |  |  |  |
| non-user vs user | 0.34 | 0.24–0.47 | 66% | <0.001 | 0.32 | 0.22–0.44 | 68% | <0.001 | 1.00 | 0.60–1.56 | 0% | 0.457 |
| non-user vs irregular | 0.67 | 0.45–0.98 | 33% | 0.018 | 0.49 | 0.31–0.74 | 51% | <0.001 | 0.80 | 0.42–1.37 | 20% | 0.113 |
| non-user vs regular | 0.08 | 0.00–0.23 | 92% | <0.001 | 0.14 | 0.00–0.38 | 86% | <0.001 | Inf | Inf–Inf | -Inf% | 1.000 |
| irregular vs regular | 0.13 | 0.00–0.37 | 87% | <0.001 | 0.28 | 0.00–0.79 | 72% | 0.070 | Inf | Inf–Inf | -Inf% | 1.000 |

## Discussion

### Re-analysis of KC22 and KB22 shows no benefit from ivermectin use on infections, hospitalisations, or death

A comparison between the key findings in KC22 and KB22 and our re-analysis is found in Table 4. The apparent risk reduction from documented artefacts — including well-known biases and sampling errors — accounts for all of the reported benefits of ivermectin claimed in both KC22 and KB22. As the key findings from Table 4 underlie all subsequent analyses in KC22 and KB22, none of the results reported in either paper holds up to scrutiny.

### The errors in KC22 and KB22 are pervasive. Should they have been obvious?

The biases and failings in KC22 and KB22 originate from a mix of sources, as outlined in Fig. 4. However, all of the issues except for missing data late in the study period (Fig. 4G) are a consequence of the study design itself. In addition, all of the issues with the possible exception of excluding private hospitalisations (Fig. 4H) lead to fictitious benefit to the ivermectin user group.

**Figure 4.**
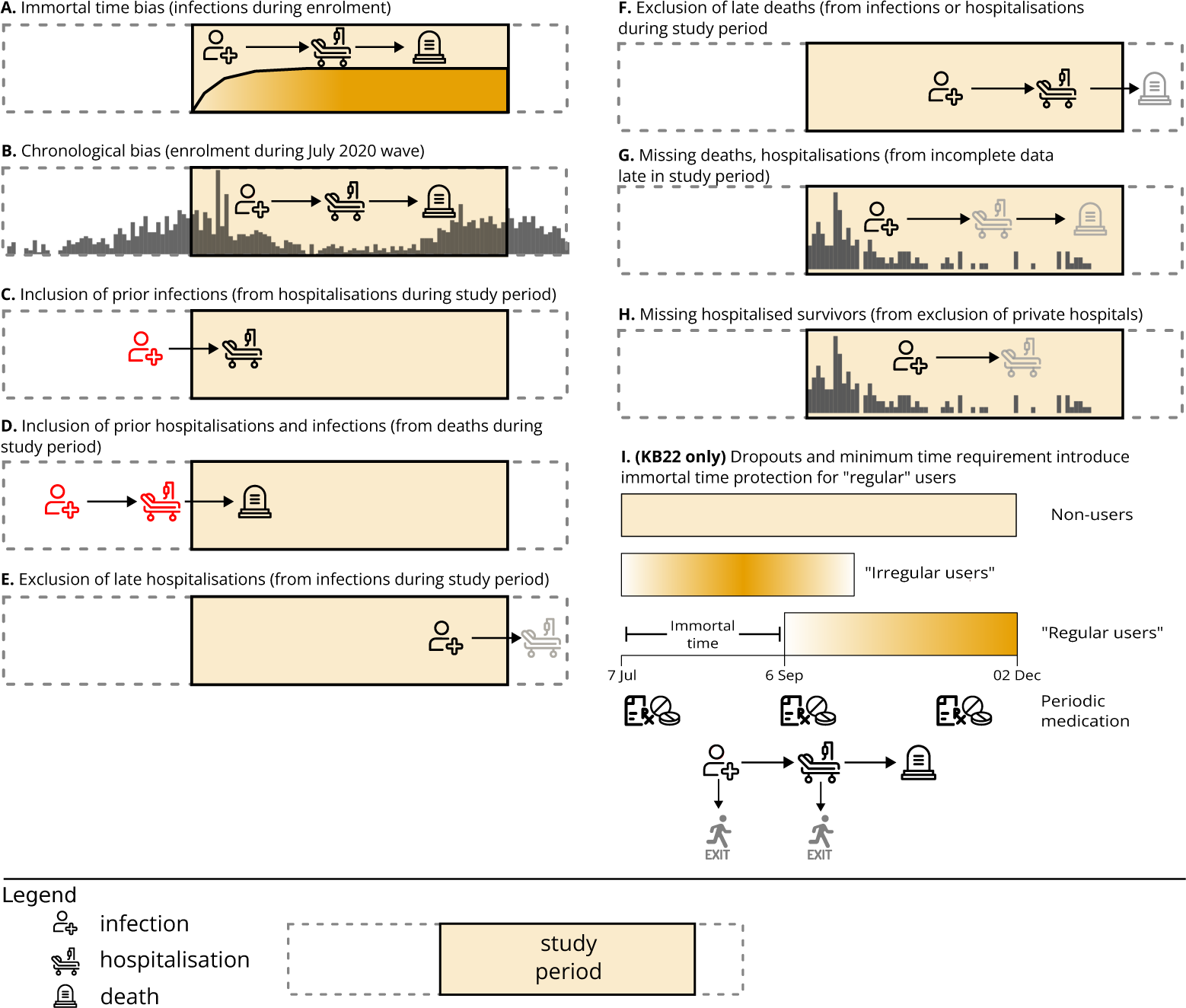
Schematic overview of different biases and errors in the design and analysis of KC22 and KB22. Red icons represent mistaken inclusion of events; gray icons represent missing, dropped, or wrongly excluded events. A-H. Biases and errors in KC22. I Additional biases and errors in KB22 (which also includes all issues in KC22).

While each issue can be considered independently, it is important to note that many of them also interact. For instance, excluding late hospitalisations and deaths (Fig. 4E-F) does not on its own benefit the ivermectin user group, but enrolment bias (Fig. 4A-B) shifts the distribution of non-user infections earlier than user infections, so attrition bias from late events (Fig. 4E-F) provides more apparent protection to users than non-users against hospitalisation and death. All of these biases should be evident, immediately or after some thought, to any scientific or medical professional with training and experience in clinical study design.

Understanding other sources of bias in KC22 and KB22 requires data. To appreciate the chronological bias during enrolment, for example, one must at least be aware of public data such as reported case rates over time. Still other issues, such as potential missing data, require careful examination, cross-tabulation, and analysis of available data to identify the extent of particular issues.

Another striking bias in KC22 and KB22, not addressed by our analysis, is the complete lack of attention to health inequity. This is hard to fathom given the likelihood of socioeconomic differences between participants and non-participants. Participation in the Itajaí program required individuals to take proactive steps: they needed to travel to distribution centres, sign up for the program, register their information, receive medication, and return periodically for medication refills. KC22 and KB22 included known *prognostic* factors in propensity score matching after infection. However, neither KC22 nor KB22 accounted for any socioeconomic factors as potential covariates affecting either infection risk or prognosis, even though impoverished and vulnerable populations are known to have higher risk and worse outcomes for many diseases and to be harder to reach and recruit for clinical studies [20, 21]. These well-known inequities were magnified during the early phases of COVID-19 because of stresses in public health systems [22–25]. Furthermore, health inequity — including during the COVID-19 pandemic — is an area of active study in Brazil, with greater risk of hospitalisation and death from social inequity and disadvantage [26–28]. Indeed, health inequity is an unavoidable concern for these study if for no other reason than the fact that hospitalisations are only reported for public hospitals (Fig. 4H).

In our view, the issues above should have been recognised and addressed by the authors, and most of them should have been recognised and questioned by the reviewers.

They were not. Instead, each paper was submitted, revised, and accepted in a matter of days, and immediately entered public discussion with a primary focus on the large size of the study population and the large magnitude of reported risk reduction, as if a large reported risk reduction must be true if the study itself is large. Unfortunately the opposite may hold if a study is built on a fallacious design. In such a case both the reported treatment effect and the apparent confidence (especially when reported as “statistical significance”) may increase with a larger study population [29–31], even if there is no actual effect of treatment. This appears to be the situation with KC22 and KB22.

### The growing consensus and persistent divide on use of ivermectin for Covid-19

Ivermectin has been known to have different mechanisms in vertebrates and nematodes for over 30 years [32]. Ivermectin’s mechanism and safety as an antiparasitic stems from its potent targeting of glutamate-gated chloride channel receptors essential for nematodes but not found in vertebrates [33]. When ivermectin was first suggested for COVID-19 based on *in vitro* experiments [4], the suggestion was not altogether unreasonable: some of the same researchers had reported a possible antiviral mechanism through the nuclear α/β1 importin complex[34, 35]. However, those studies were undertaken in cell lines such as Vero E6 or HeLa (both widely used for viral assays); when careful comparison studies were carried out in more relevant human bronchial epithelial cells, ivermectin had no effect on SARS-CoV-2 replication [36]. There were other reasons to be sceptical of the initial enthusiasm; the well-studied pharmacokinetics and pharmacodynamics of ivermectin strongly imply that it would be impossible to replicate in humans the concentrations required for *in vitro* activity [37, 38].

Some doctors began prescribing ivermectin based on preliminary and preclinical studies, even as most mainstream public health authorities encouraged waiting for clinical trials. As a result, hundreds of studies of ivermectin for COVID-19 have been published in the last three years, including dozens of clinical trials, numerous cell biology and biochemical studies, mechanistic speculation based on molecular docking, and competing reviews. Some of the earliest high-profile studies reporting large effects of ivermectin have been retracted or otherwise flagged for ethical concerns [39, 40]. Meta-analyses that included such flawed analysis have also been retracted [41] and reviews advocating for the immediate use of ivermectin [42] have been criticised as deeply problematic [43]. One possible lesson from both ivermectin metaanalysis and vaccine clinical trials is to require use of anonymised individual patient data, rather than summary data alone, for assessment of bias in meta-analysis [44, 45].

Recent rigorous randomised clinical trials have largely not found clinical benefit for ivermectin use in COVID-19 [46–50], strengthening the growing scientific and clinical consensus not to recommend clinical use of ivermectin for COVID-19 treatment or prophylaxis [51]. This consensus remains persistently rejected by ivermectin advocates citing smaller RCTs or observational studies that appear to show an enormous effect of ivermectin. As with any area of active research, interpretations regarding ivermectin’s efficacy might differ based on the chosen data sets and their assessments, underscoring the need for more thorough and objective analysis of the available literature. What began as a scientific disagreement has become a challenge for effective science communication in the context of public health.

This work focuses on the fallacies in KB22 and KC22 specifically, rather than any general question of ivermectin for COVID-19. Readers interested in that question should refer to [51]. Our work highlights the dangers posed by scientific misinformation. Not only have the unfounded conclusions of these papers been used to spread misinformation, methodological criticism of them leads to aggressive and misleading attacks, as happened upon the initial release of this study as a preprint [52, 53]. Regional, national, and international organisations^1^ have sprung up to advocate for ivermectin and other non-proven or weakly supported treatments for COVID-19, and strive to influence public opinion. Ivermectin-for-COVID advocacy groups maintain their position and influence through mechanisms including promoting “science by preprint”, exploiting perfunctory peer review, aggressively using social media, and cultivating socio-political alliances including the anti-vaccination movement. Observational studies are particularly vulnerable to misinterpretation and use as misinformation.

### The use of simulation to solve statistical fallacies

Statistical fallacies or ‘statistical lies’ affect our lives in many ways: we read them in newspapers, we hear them in conversation, we inadvertently make them ourselves, and they are unfortunately common in science. The fallacies often go hand-in-hand with cognitive heuristics that bias our perception of reality [54]. For instance, salience bias (the tendency to focus on remarkable events or prominent features) leads people to overestimate risk of rare events and make decisions that appear irrational and incur a cost to themselves and to society [55, 56].

Despite the review process, biases and statistical fallacies also arise in scientific literature [19, 57], and proliferation of these fallacies carries the potential for disaster: the incorrect conclusions of KC22 and KB22 for instance, were used to support arguments that ivermectin was at least as effective as vaccination against COVID-19 related death, potentially increasing vaccine hesitancy and thereby increasing the global death toll due to COVID-19.

One of the most famous statistical fallacies is seen in the Monty Hall problem, named for the original host of the American television show “Let’s Make a Deal”, where a variant of this puzzle appeared in every episode. In this puzzle, a player is presented three closed doors and asked to choose one. Behind one of these doors is a prize which the player will win if they choose the correct door. Once the player has chosen a door, the host reveals which of the other two doors does not contain the prize, and subsequently asks the player if they would like to stick with their initial choice or switch to the other closed door. When presented for the first time, most people assume that switching their choice will not affect the likelihood of winning a prize [58] The answer, however^2^ is that the likelihood of winning a prize after switching is two thirds, whereas the likelihood of winning is only one third when the participant doesn’t switch doors.

Remarkably, even when people are shown explanations, simulations and mathematical proofs, many - including renown statisticians - still refuse to accept the answer of the puzzle [58, 59]. Studies using repeated simulations of the Monte Hall problem show a remarkable adoption of the correct answer. Herein participants play the game over and over on the computer, and get feedback on how often they won the prize. The Monte Carlo simulation that we use here is basically an automation of this process. Akin to the Monte Hall problem, researchers may falsely reason that it is correct to include participants that got infected by COVID-19 before registering/consuming ivermectin as non-users in a rolling registration context. But simulating the process unveils that an artificial efficacy emerges for the treatment group. We hope therefore that our work extends beyond a correction of these two papers, and helps researchers of observational trials in general not to repeat these fallacies.

## Methods and materials

### Starting data

We used data made available by the authors of KC22 at DOI 10.17605/OSF.IO/UXHAF. This data was missing critical information, which we addressed as follows. City-wide monthly infections and deaths were taken from [43], which in turn obtained them from the Brazilian Health Ministry. We also used the Brazilian Health Ministry resource to obtain national reporting data for individuals including dates of symptom onset, hospitalisation, and death, though this was largely limited to hospitalised patients during the time of the study. We downloaded this data on 26 September 2022. While the study began enrolling participants on July 7, 2020, the KC22 data does not indicate when any individual patient joined the program and was provided ivermectin. We used an estimate of program enrolment over time by following municipal bulletins from Itajaí as well as contemporaneous articles in the local Itajaí press (e.g. [60]). Sources for each estimate are in the GitHub repository.

### Analysis of overlap between KC22 and Brazilian Health Ministry data

Data from KC22 was cross-tabulated with public data from the Brazilian Health Ministry. After identifying variables that were available in both data sets, we identified matches between infected individuals (both users and non-users) from KC22 and infected individuals from the health ministry data. Matches were counted only if they were identical for all of the variables considered (birth date, hospitalisation status, sex, and death outcome). We considered records of deaths and hospitalised survivors separately. When matching on deaths, we ignored hospitalisation because hospitalisation details were apparently incorrect for many records of deceased individuals in the KC22 data. Specifically, 95% of death records in the Itajaí citywide data from DATASUS were marked as hospitalised, but only 65% of deaths in the KC22 data were marked as hospitalised.

A few entries in KC22 had more than one match to the health ministry data, which could be due to data errors or multiple infections. We included only the single match with the latest date of symptom onset in order to avoid over-counting pre-study inclusion in KC22 and to minimise the estimate of immortal time bias.

### Allocation to exposure and outcome groups

For each simulation experiment, repeated Bernoulli trials were conducted for each of the 159,560 individuals to assign them to exposure groups (users or non users) as well as outcomes (infections, hospitalisations, deaths). Hospitalisation was always preceded by infection, and death was always preceded by hospitalisation. All allocation probabilities were fixed based on the number of users, non-users, infections, hospitalisations, and deaths reported in KC22 (e.g. probability to be an ivermectin user is 113,844 / 159,560).

### Simulation of event times and ivermectin usage

For each simulated individual that contracted COVID-19, was hospitalised, or died, the event date was sampled from the empirical distributions of official government records. In the case of missing data, we used complete cases to confirm published reports that time periods in the progression of COVID-19 were well-approximated by a Weibull distribution [61–63], and then created an objective function using the empirical mean and standard deviation of time delays to numerically optimise the Weibull scale and shape parameters to impute missing event dates. In the i-ENR and i-INF models, the empirical distribution of symptom onset times were used identically, but limited pre-study onset (< 1 month) was possible in the i-INF model provided the notification date was within the study period. In the case of the i-KC22 model, we sampled onset dates for matched records for those marked as hospitalised or deceased. For those not marked as hospitalised or deceased, imputed onset dates from a smooth (loess) distribution of onset dates from the matched records. We included unmatched late (post-study period) hospitalisations and deaths in order to estimate the impact of these biases in Table 3.

Intended ivermectin usage was truncated to actual usage as follows. In probabilistic truncation, infected users with high intended usage (≥30 pills) were less likely (p = 0.05) to stop usage on infection than those with lower intended usage (p = 0.3). In deterministic truncation, all infected users stopped ivermectin use upon infection. In all cases, infected users stopped upon hospitalisation. Ivermectin users where split into irregular (≤10 pills) and regular users (≥30 pills) based on simulated actual usage.

### Calculation of simulated estimates

Risk ratios were calculated using unconditional maximum likelihood (Wald statistic), while confidence intervals and p values estimated from 1000 bootstraps using the riskratio.boot function from the epitools package in R [64]. Summary statistics from KC22 and KB22 were reestimated the same way for consistency. For summary statistics after correction, we assumed any true effects of ivermectin were independent of apparent effects from artefacts and errors. Risk ratios, confidence intervals, and p values after correction were calculated by 1000 bootstraps of simulated and reported data for each simulation.

## Data and code availability

All data is publicly available from previous publications or public databases of the Brazil Ministry of Health. Links are provided at the end of the Methods section. Source code to reproduce the analyses here is available at https://github.com/gtuckerkellogg/itajai-reanalysis.

## Ethics approval

The Institutional Review Board of the National University of Singapore waived ethical approval of this work based on sole use of previously approved and publicly available subject data. Reference NUS-IRB-2023-474.

## Author contributions

RM conducted the research required to uncover all fallacies mentioned in the manuscript (delayed registrations, missing data, incorrect inclusion of prior infections, all biases). GTK independently uncovered and estimated the immortal time bias, enrolment bias, and attrition bias. RM and GTK wrote the simulation and analysis code. GTK in particular, RM, and ACPA contributed to writing and reviewing the manuscript. ACPA acquired the data from the Brazilian Health Ministry. ACPA has been in frequent contact with Itajaí City Hall in an attempt to get access to missing raw data. RM has been in frequent contact with the KC22 and KB22 authors in an attempt to discuss the issues in their work before publishing this manuscript. RM has additionally been in frequent contact with the authors following the initial preprint release, attempting to address the incorrect public statements made by the authors of KC22 and KB22.

## Acknowledgements

We thank Gideon Meyerowitz-Katz, Lisa Tucker-Kellogg, Andrew Gelman, Amichai Perlman, and Annelot Mills for their valuable comments and feedback. We would like to thank Daniel Victor Tausk for identifying issues related to DATASUS filtering in our preprint and for recommending more precise methods to match DATASUS with data from KC22.

## Footnotes

1 These include the Front Line COVID-19 Critical Care Alliance (FLCCC) and America’s Frontline Doctors in the United States, Médicos Pela Vida in Brazil, and the World Council for Health.

2 in the simplest case, assuming that the host always opens the wrong door and always gives the player a choice to switch

