## Supplemental tables and figures for "Published benefits of ivermectin use in Itajaí, Brazil for COVID-19 infection, hospitalisation, and mortality are entirely explained by statistical artefacts"

### List of Tables

S1 i-ENR model with probabilistic stop on infection. Stop probability: 0.30 (irregular), 0.05 (regular). Statistics for hospitalisations and deaths are reported for all individuals in the cohort. . . . 3

S2 i-ENR model with uniform stop on infection. Stop probability: 1.0 (all individuals). Statistics for hospitalisations and deaths are reported for all individuals in the cohort. 4

S3 i-ENR model with probabilistic stop on infection. Stop probability: 0.30 (irregular), 0.05 (regular). Statistics for hospitalisations and deaths are limited to infected individuals. . . . 4

S4 i-ENR model with uniform stop on infection. Stop probability: 1.0 (all individuals). Statistics for hospitalisations and deaths are limited to infected individuals. . . . 5

S5 i-INF model with probabilistic stop on infection. Stop probability: 0.30 (irregular), 0.05 (regular). Statistics for hospitalisations and deaths are reported for all individuals in the cohort. . . . 6

S6 i-INF model with uniform stop on infection. Stop probability: 1.0 (all individuals). Statistics for hospitalisations and deaths are reported for all individuals in the cohort. 7

S7 i-INF model with probabilistic stop on infection. Stop probability: 0.30 (irregular), 0.05 (regular). Statistics for hospitalisations and deaths are limited to infected individuals. . . . 7

S8 i-INF model with uniform stop on infection. Stop probability: 1.0 (all individuals). Statistics for hospitalisations and deaths are limited to infected individuals. . . . 8

S9 i-KC22 model with probabilistic stop on infection. Stop probability: 0.30 (irregular), 0.05 (regular). Statistics for hospitalisations and deaths are reported for all individuals in the cohort. . . . 9

S10 i-KC22 model with uniform stop on infection. Stop probability: 1.0 (all individuals). Statistics for hospitalisations and deaths are reported for all individuals in the cohort. 10

S11 i-KC22 model with probabilistic stop on infection. Stop probability: 0.30 (irregular), 0.05 (regular). Statistics for hospitalisations and deaths are limited to infected individuals. . . . 10

S12 i-KC22 model with uniform stop on infection. Stop probability: 1.0 (all individuals). Statistics for hospitalisations and deaths are limited to infected individuals. . . . 11

### List of Figures

S1 "Missed" deaths . . . 12
S2 Attrition bias in the i-INF model . . . 13
S3 Attrition bias in the i-KC22 model . . . 14
S4 Apparent protection of "regular" users in the i-INF model . . . 15
S5 Apparent protection of "regular" users in the i-KC22 model . . . 16

### i-ENR model simulations

| | Simulation | | | | KB22/KC22 | | | |
|---|---|---|---|---|---|---|---|---|
| Summary statistics | RR | 95% CI | Risk Red. | p.val | RR | 95% CI | Risk Red. | p.val |
| **Infection** | | | | | | | | |
| non-user vs user | 0.83 | 0.80–0.88 | 17% | <0.001 | 0.56 | 0.53–0.58 | 44% | <0.001 |
| non-user vs irregular | 0.94 | 0.88–1.00 | 6% | 0.022 | 0.68 | 0.64–0.73 | 32% | <0.001 |
| non-user vs regular | 0.72 | 0.64–0.81 | 28% | <0.001 | 0.51 | 0.45–0.57 | 49% | <0.001 |
| irregular vs regular | 0.77 | 0.68–0.86 | 23% | <0.001 | 0.75 | 0.66–0.84 | 25% | <0.001 |
| **Hospitalisation** | | | | | | | | |
| non-user vs user | 0.75 | 0.56–1.03 | 25% | 0.039 | 0.35 | 0.26–0.47 | 65% | <0.001 |
| non-user vs irregular | 1.23 | 0.86–1.76 | -23% | 0.856 | 0.52 | 0.34–0.74 | 48% | <0.001 |
| non-user vs regular | 0.33 | 0.06–0.72 | 67% | 0.002 | 0.00 | 0.00–0.00 | 100% | <0.001 |
| irregular vs regular | 0.27 | 0.05–0.60 | 73% | <0.001 | 0.00 | 0.00–0.00 | 100% | <0.001 |
| **Death** | | | | | | | | |
| non-user vs user | 0.71 | 0.50–1.02 | 29% | 0.026 | 0.32 | 0.23–0.44 | 68% | <0.001 |
| non-user vs irregular | 1.26 | 0.84–1.89 | -26% | 0.837 | 0.49 | 0.31–0.74 | 51% | <0.001 |
| non-user vs regular | 0.22 | 0.02–0.57 | 78% | 0.002 | 0.14 | 0.00–0.38 | 86% | <0.001 |
| irregular vs regular | 0.18 | 0.01–0.46 | 82% | <0.001 | 0.28 | 0.00–0.82 | 72% | 0.070 |

**Table S1**
i-ENR model with probabilistic stop on infection. Stop probability: 0.30 (irregular), 0.05 (regular). Statistics for hospitalisations and deaths are reported for all individuals in the cohort.

| Summary statistics | Simulation | | | | KB22/KC22 | | | |
|---|---|---|---|---|---|---|---|---|
| | RR | 95% CI | Risk Red. | p.val | RR | 95% CI | Risk Red. | p.val |
| **Infection** | | | | | | | | |
| non-user vs user | 0.83 | 0.80–0.88 | 17% | <0.001 | 0.56 | 0.53–0.58 | 44% | <0.001 |
| non-user vs irregular | 1.18 | 1.11–1.25 | -18% | 1.000 | 0.68 | 0.64–0.72 | 32% | <0.001 |
| non-user vs regular | 0.45 | 0.39–0.52 | 55% | <0.001 | 0.51 | 0.45–0.58 | 49% | <0.001 |
| irregular vs regular | 0.38 | 0.33–0.44 | 62% | <0.001 | 0.75 | 0.66–0.85 | 25% | <0.001 |
| **Hospitalisation** | | | | | | | | |
| non-user vs user | 0.75 | 0.56–1.03 | 25% | 0.039 | 0.35 | 0.26–0.46 | 65% | <0.001 |
| non-user vs irregular | 1.30 | 0.92–1.86 | -30% | 0.915 | 0.52 | 0.35–0.73 | 48% | <0.001 |
| non-user vs regular | 0.32 | 0.06–0.71 | 68% | 0.002 | 0.00 | 0.00–0.00 | 100% | <0.001 |
| irregular vs regular | 0.25 | 0.04–0.56 | 75% | <0.001 | 0.00 | 0.00–0.00 | 100% | <0.001 |
| **Death** | | | | | | | | |
| non-user vs user | 0.71 | 0.50–1.02 | 29% | 0.026 | 0.32 | 0.22–0.44 | 68% | <0.001 |
| non-user vs irregular | 1.35 | 0.91–2.01 | -35% | 0.926 | 0.49 | 0.32–0.74 | 51% | <0.001 |
| non-user vs regular | 0.21 | 0.01–0.55 | 79% | 0.001 | 0.14 | 0.00–0.38 | 86% | <0.001 |
| irregular vs regular | 0.16 | 0.01–0.41 | 84% | <0.001 | 0.28 | 0.00–0.78 | 72% | 0.070 |

**Table S2**

i-ENR model with uniform stop on infection. Stop probability: 1.0 (all individuals). Statistics for hospitalisations and deaths are reported for all individuals in the cohort.

| Summary statistics | Simulation | | | | KB22/KC22 | | | |
|---|---|---|---|---|---|---|---|---|
| | RR | 95% CI | Risk Red. | p.val | RR | 95% CI | Risk Red. | p.val |
| **Infection** | | | | | | | | |
| non-user vs user | 0.83 | 0.80–0.88 | 17% | <0.001 | 0.56 | 0.53–0.58 | 44% | <0.001 |
| non-user vs irregular | 0.94 | 0.88–1.00 | 6% | 0.022 | 0.68 | 0.64–0.72 | 32% | <0.001 |
| non-user vs regular | 0.72 | 0.64–0.81 | 28% | <0.001 | 0.51 | 0.45–0.58 | 49% | <0.001 |
| irregular vs regular | 0.77 | 0.68–0.86 | 23% | <0.001 | 0.75 | 0.66–0.85 | 25% | <0.001 |
| **Hospitalisation** | | | | | | | | |
| non-user vs user | 0.90 | 0.67–1.23 | 10% | 0.217 | 0.63 | 0.47–0.83 | 37% | 0.001 |
| non-user vs irregular | 1.31 | 0.92–1.86 | -31% | 0.916 | 0.76 | 0.51–1.08 | 24% | 0.143 |
| non-user vs regular | 0.45 | 0.09–1.00 | 55% | 0.034 | 0.00 | 0.00–0.00 | 100% | <0.001 |
| irregular vs regular | 0.35 | 0.07–0.78 | 65% | 0.001 | 0.00 | 0.00–0.00 | 100% | 0.003 |
| **Death** | | | | | | | | |
| non-user vs user | 0.85 | 0.61–1.22 | 15% | 0.150 | 0.57 | 0.40–0.80 | 43% | <0.001 |
| non-user vs irregular | 1.34 | 0.90–2.00 | -34% | 0.907 | 0.72 | 0.45–1.07 | 28% | 0.149 |
| non-user vs regular | 0.31 | 0.02–0.78 | 69% | 0.005 | 0.27 | 0.00–0.74 | 73% | 0.044 |
| irregular vs regular | 0.23 | 0.02–0.60 | 77% | <0.001 | 0.38 | 0.00–1.09 | 62% | 0.212 |

**Table S3**

i-ENR model with probabilistic stop on infection. Stop probability: 0.30 (irregular), 0.05 (regular). Statistics for hospitalisations and deaths are limited to infected individuals.

| | Simulation | | | | KB22/KC22 | | | |
|---|---|---|---|---|---|---|---|---|
| Summary statistics | RR | 95% CI | Risk Red. | p.val | RR | 95% CI | Risk Red. | p.val |
| **Infection** | | | | | | | | |
| non-user vs user | 0.83 | 0.80–0.88 | 17% | <0.001 | 0.56 | 0.53–0.58 | 44% | <0.001 |
| non-user vs irregular | 1.18 | 1.11–1.25 | -18% | 1.000 | 0.68 | 0.65–0.73 | 32% | <0.001 |
| non-user vs regular | 0.45 | 0.39–0.52 | 55% | <0.001 | 0.51 | 0.45–0.58 | 49% | <0.001 |
| irregular vs regular | 0.38 | 0.33–0.44 | 62% | <0.001 | 0.75 | 0.66–0.84 | 25% | <0.001 |
| **Hospitalisation** | | | | | | | | |
| non-user vs user | 0.90 | 0.67–1.23 | 10% | 0.217 | 0.63 | 0.47–0.83 | 37% | 0.001 |
| non-user vs irregular | 1.11 | 0.78–1.56 | -11% | 0.687 | 0.76 | 0.50–1.07 | 24% | 0.143 |
| non-user vs regular | 0.71 | 0.13–1.57 | 29% | 0.215 | 0.00 | 0.00–0.00 | 100% | <0.001 |
| irregular vs regular | 0.65 | 0.12–1.44 | 35% | 0.168 | 0.00 | 0.00–0.00 | 100% | 0.003 |
| **Death** | | | | | | | | |
| non-user vs user | 0.85 | 0.61–1.22 | 15% | 0.150 | 0.57 | 0.41–0.79 | 43% | <0.001 |
| non-user vs irregular | 1.15 | 0.78–1.69 | -15% | 0.723 | 0.72 | 0.45–1.07 | 28% | 0.149 |
| non-user vs regular | 0.46 | 0.03–1.20 | 54% | 0.084 | 0.27 | 0.00–0.71 | 73% | 0.044 |
| irregular vs regular | 0.41 | 0.03–1.07 | 59% | 0.046 | 0.38 | 0.00–1.09 | 62% | 0.212 |

**Table S4**

i-ENR model with uniform stop on infection. Stop probability: 1.0 (all individuals). Statistics for hospitalisations and deaths are limited to infected individuals.

### i-INF model simulations

| | Simulation | | | | KB22/KC22 | | | |
|---|---|---|---|---|---|---|---|---|
| Summary statistics | RR | 95% CI | Risk Red. | p.val | RR | 95% CI | Risk Red. | p.val |
| **Infection** | | | | | | | | |
| non-user vs user | 0.73 | 0.70–0.76 | 27% | <0.001 | 0.56 | 0.53–0.58 | 44% | <0.001 |
| non-user vs irregular | 0.82 | 0.77–0.87 | 18% | <0.001 | 0.68 | 0.64–0.72 | 32% | <0.001 |
| non-user vs regular | 0.63 | 0.55–0.70 | 37% | <0.001 | 0.51 | 0.45–0.57 | 49% | <0.001 |
| irregular vs regular | 0.76 | 0.67–0.86 | 24% | <0.001 | 0.75 | 0.66–0.84 | 25% | <0.001 |
| **Hospitalisation** | | | | | | | | |
| non-user vs user | 0.59 | 0.44–0.80 | 41% | <0.001 | 0.35 | 0.26–0.47 | 65% | <0.001 |
| non-user vs irregular | 0.97 | 0.67–1.37 | 3% | 0.380 | 0.52 | 0.35–0.74 | 48% | <0.001 |
| non-user vs regular | 0.25 | 0.04–0.56 | 75% | <0.001 | 0.00 | 0.00–0.00 | 100% | <0.001 |
| irregular vs regular | 0.26 | 0.04–0.61 | 74% | 0.001 | 0.00 | 0.00–0.00 | 100% | <0.001 |
| **Death** | | | | | | | | |
| non-user vs user | 0.54 | 0.39–0.77 | 46% | 0.002 | 0.32 | 0.22–0.44 | 68% | <0.001 |
| non-user vs irregular | 0.97 | 0.64–1.43 | 3% | 0.384 | 0.49 | 0.31–0.75 | 51% | <0.001 |
| non-user vs regular | 0.16 | 0.01–0.42 | 84% | <0.001 | 0.14 | 0.00–0.39 | 86% | <0.001 |
| irregular vs regular | 0.17 | 0.01–0.46 | 83% | <0.001 | 0.28 | 0.00–0.82 | 72% | 0.070 |

**Table S5**
i-INF model with probabilistic stop on infection. Stop probability: 0.30 (irregular), 0.05 (regular). Statistics for hospitalisations and deaths are reported for all individuals in the cohort.

| Summary statistics | Simulation | | | | KB22/KC22 | | | |
|---|---|---|---|---|---|---|---|---|
| | RR | 95% CI | Risk Red. | p.val | RR | 95% CI | Risk Red. | p.val |
| **Infection** | | | | | | | | |
| non-user vs user | 0.73 | 0.70–0.76 | 27% | <0.001 | 0.56 | 0.53–0.58 | 44% | <0.001 |
| non-user vs irregular | 1.03 | 0.98–1.09 | -3% | 0.867 | 0.68 | 0.64–0.73 | 32% | <0.001 |
| non-user vs regular | 0.39 | 0.33–0.45 | 61% | <0.001 | 0.51 | 0.45–0.58 | 49% | <0.001 |
| irregular vs regular | 0.38 | 0.32–0.44 | 62% | <0.001 | 0.75 | 0.66–0.85 | 25% | <0.001 |
| **Hospitalisation** | | | | | | | | |
| non-user vs user | 0.59 | 0.44–0.80 | 41% | <0.001 | 0.35 | 0.26–0.46 | 65% | <0.001 |
| non-user vs irregular | 1.02 | 0.72–1.44 | -2% | 0.509 | 0.52 | 0.35–0.74 | 48% | <0.001 |
| non-user vs regular | 0.24 | 0.04–0.56 | 76% | <0.001 | 0.00 | 0.00–0.00 | 100% | <0.001 |
| irregular vs regular | 0.24 | 0.04–0.56 | 76% | <0.001 | 0.00 | 0.00–0.00 | 100% | <0.001 |
| **Death** | | | | | | | | |
| non-user vs user | 0.54 | 0.39–0.77 | 46% | 0.002 | 0.32 | 0.22–0.45 | 68% | <0.001 |
| non-user vs irregular | 1.04 | 0.70–1.52 | -4% | 0.527 | 0.49 | 0.31–0.74 | 51% | <0.001 |
| non-user vs regular | 0.15 | 0.01–0.40 | 85% | <0.001 | 0.14 | 0.00–0.38 | 86% | <0.001 |
| irregular vs regular | 0.15 | 0.01–0.41 | 85% | <0.001 | 0.28 | 0.00–0.82 | 72% | 0.070 |

**Table S6**
i-INF model with uniform stop on infection. Stop probability: 1.0 (all individuals). Statistics for hospitalisations and deaths are reported for all individuals in the cohort.

| Summary statistics | Simulation | | | | KB22/KC22 | | | |
|---|---|---|---|---|---|---|---|---|
| | RR | 95% CI | Risk Red. | p.val | RR | 95% CI | Risk Red. | p.val |
| **Infection** | | | | | | | | |
| non-user vs user | 0.73 | 0.70–0.76 | 27% | <0.001 | 0.56 | 0.53–0.58 | 44% | <0.001 |
| non-user vs irregular | 0.82 | 0.77–0.87 | 18% | <0.001 | 0.68 | 0.64–0.73 | 32% | <0.001 |
| non-user vs regular | 0.63 | 0.55–0.70 | 37% | <0.001 | 0.51 | 0.45–0.57 | 49% | <0.001 |
| irregular vs regular | 0.76 | 0.67–0.86 | 24% | <0.001 | 0.75 | 0.66–0.85 | 25% | <0.001 |
| **Hospitalisation** | | | | | | | | |
| non-user vs user | 0.81 | 0.61–1.09 | 19% | 0.066 | 0.63 | 0.47–0.83 | 37% | 0.001 |
| non-user vs irregular | 1.18 | 0.82–1.65 | -18% | 0.798 | 0.76 | 0.51–1.07 | 24% | 0.143 |
| non-user vs regular | 0.40 | 0.06–0.89 | 60% | 0.018 | 0.00 | 0.00–0.00 | 100% | <0.001 |
| irregular vs regular | 0.35 | 0.06–0.79 | 65% | 0.005 | 0.00 | 0.00–0.00 | 100% | 0.003 |
| **Death** | | | | | | | | |
| non-user vs user | 0.74 | 0.53–1.05 | 26% | 0.040 | 0.57 | 0.40–0.79 | 43% | <0.001 |
| non-user vs irregular | 1.17 | 0.79–1.73 | -17% | 0.766 | 0.72 | 0.46–1.07 | 28% | 0.149 |
| non-user vs regular | 0.26 | 0.01–0.67 | 74% | 0.003 | 0.27 | 0.00–0.71 | 73% | 0.044 |
| irregular vs regular | 0.22 | 0.01–0.60 | 78% | 0.002 | 0.38 | 0.00–1.05 | 62% | 0.212 |

**Table S7**
i-INF model with probabilistic stop on infection. Stop probability: 0.30 (irregular), 0.05 (regular). Statistics for hospitalisations and deaths are limited to infected individuals.

| | Simulation | | | | KB22/KC22 | | | |
|---|---|---|---|---|---|---|---|---|
| Summary statistics | RR | 95% CI | Risk Red. | p.val | RR | 95% CI | Risk Red. | p.val |
| **Infection** | | | | | | | | |
| non-user vs user | 0.73 | 0.70–0.76 | 27% | <0.001 | 0.56 | 0.53–0.58 | 44% | <0.001 |
| non-user vs irregular | 1.03 | 0.98–1.09 | -3% | 0.867 | 0.68 | 0.65–0.73 | 32% | <0.001 |
| non-user vs regular | 0.39 | 0.33–0.45 | 61% | <0.001 | 0.51 | 0.45–0.58 | 49% | <0.001 |
| irregular vs regular | 0.38 | 0.32–0.44 | 62% | <0.001 | 0.75 | 0.66–0.85 | 25% | <0.001 |
| **Hospitalisation** | | | | | | | | |
| non-user vs user | 0.81 | 0.61–1.09 | 19% | 0.066 | 0.63 | 0.47–0.83 | 37% | 0.001 |
| non-user vs irregular | 0.99 | 0.70–1.39 | 1% | 0.436 | 0.76 | 0.51–1.08 | 24% | 0.143 |
| non-user vs regular | 0.62 | 0.10–1.41 | 38% | 0.147 | 0.00 | 0.00–0.00 | 100% | <0.001 |
| irregular vs regular | 0.64 | 0.10–1.46 | 36% | 0.153 | 0.00 | 0.00–0.00 | 100% | 0.003 |
| **Death** | | | | | | | | |
| non-user vs user | 0.74 | 0.53–1.05 | 26% | 0.040 | 0.57 | 0.41–0.79 | 43% | <0.001 |
| non-user vs irregular | 1.00 | 0.68–1.46 | -0% | 0.464 | 0.72 | 0.45–1.08 | 28% | 0.149 |
| non-user vs regular | 0.38 | 0.02–1.02 | 62% | 0.041 | 0.27 | 0.00–0.74 | 73% | 0.044 |
| irregular vs regular | 0.39 | 0.02–1.06 | 61% | 0.047 | 0.38 | 0.00–1.05 | 62% | 0.212 |

**Table S8**
i-INF model with uniform stop on infection. Stop probability: 1.0 (all individuals). Statistics for hospitalisations and deaths are limited to infected individuals.

### i-KC22 model simulations

| | Simulation | | | | KB22/KC22 | | | |
|---|---|---|---|---|---|---|---|---|
| Summary statistics | RR | 95% CI | Risk Red. | p.val | RR | 95% CI | Risk Red. | p.val |
| **Infection** | | | | | | | | |
| non-user vs user | 0.49 | 0.47–0.51 | 51% | <0.001 | 0.56 | 0.53–0.58 | 44% | <0.001 |
| non-user vs irregular | 0.64 | 0.60–0.68 | 36% | <0.001 | 0.68 | 0.64–0.73 | 32% | <0.001 |
| non-user vs regular | 0.37 | 0.33–0.43 | 63% | <0.001 | 0.51 | 0.45–0.57 | 49% | <0.001 |
| irregular vs regular | 0.59 | 0.51–0.67 | 41% | <0.001 | 0.75 | 0.66–0.85 | 25% | <0.001 |
| **Hospitalisation** | | | | | | | | |
| non-user vs user | 0.40 | 0.28–0.56 | 60% | <0.001 | 0.35 | 0.26–0.47 | 65% | <0.001 |
| non-user vs irregular | 0.74 | 0.49–1.09 | 26% | 0.045 | 0.52 | 0.34–0.74 | 48% | <0.001 |
| non-user vs regular | 0.15 | 0.01–0.39 | 85% | <0.001 | 0.00 | 0.00–0.00 | 100% | <0.001 |
| irregular vs regular | 0.21 | 0.01–0.56 | 79% | <0.001 | 0.00 | 0.00–0.00 | 100% | <0.001 |
| **Death** | | | | | | | | |
| non-user vs user | 0.34 | 0.24–0.47 | 66% | <0.001 | 0.32 | 0.22–0.44 | 68% | <0.001 |
| non-user vs irregular | 0.67 | 0.45–0.98 | 33% | 0.018 | 0.49 | 0.31–0.74 | 51% | <0.001 |
| non-user vs regular | 0.08 | 0.00–0.23 | 92% | <0.001 | 0.14 | 0.00–0.38 | 86% | <0.001 |
| irregular vs regular | 0.13 | 0.00–0.37 | 87% | <0.001 | 0.28 | 0.00–0.79 | 72% | 0.070 |

**Table S9**
i-KC22 model with probabilistic stop on infection. Stop probability: 0.30 (irregular), 0.05 (regular). Statistics for hospitalisations and deaths are reported for all individuals in the cohort.

| Summary statistics | Simulation RR | 95% CI | Risk Red. | p.val | KB22/KC22 RR | 95% CI | Risk Red. | p.val |
|---|---|---|---|---|---|---|---|---|
| **Infection** | | | | | | | | |
| non-user vs user | 0.49 | 0.47–0.51 | 51% | <0.001 | 0.56 | 0.53–0.58 | 44% | <0.001 |
| non-user vs irregular | 1.00 | 0.95–1.05 | 0% | 0.450 | 0.68 | 0.64–0.73 | 32% | <0.001 |
| non-user vs regular | 0.10 | 0.07–0.12 | 90% | <0.001 | 0.51 | 0.45–0.58 | 49% | <0.001 |
| irregular vs regular | 0.10 | 0.07–0.12 | 90% | <0.001 | 0.75 | 0.66–0.85 | 25% | <0.001 |
| **Hospitalisation** | | | | | | | | |
| non-user vs user | 0.40 | 0.28–0.56 | 60% | <0.001 | 0.35 | 0.26–0.47 | 65% | <0.001 |
| non-user vs irregular | 0.76 | 0.51–1.11 | 24% | 0.062 | 0.52 | 0.34–0.73 | 48% | <0.001 |
| non-user vs regular | 0.14 | 0.01–0.37 | 86% | <0.001 | 0.00 | 0.00–0.00 | 100% | <0.001 |
| irregular vs regular | 0.19 | 0.01–0.52 | 81% | <0.001 | 0.00 | 0.00–0.00 | 100% | <0.001 |
| **Death** | | | | | | | | |
| non-user vs user | 0.34 | 0.24–0.47 | 66% | <0.001 | 0.32 | 0.23–0.44 | 68% | <0.001 |
| non-user vs irregular | 0.70 | 0.47–1.01 | 30% | 0.023 | 0.49 | 0.31–0.74 | 51% | <0.001 |
| non-user vs regular | 0.08 | 0.00–0.22 | 92% | <0.001 | 0.14 | 0.00–0.37 | 86% | <0.001 |
| irregular vs regular | 0.11 | 0.00–0.33 | 89% | <0.001 | 0.28 | 0.00–0.82 | 72% | 0.070 |

**Table S10**
i-KC22 model with uniform stop on infection. Stop probability: 1.0 (all individuals). Statistics for hospitalisations and deaths are reported for all individuals in the cohort.

| Summary statistics | Simulation RR | 95% CI | Risk Red. | p.val | KB22/KC22 RR | 95% CI | Risk Red. | p.val |
|---|---|---|---|---|---|---|---|---|
| **Infection** | | | | | | | | |
| non-user vs user | 0.49 | 0.47–0.51 | 51% | <0.001 | 0.56 | 0.53–0.58 | 44% | <0.001 |
| non-user vs irregular | 0.64 | 0.60–0.68 | 36% | <0.001 | 0.68 | 0.64–0.72 | 32% | <0.001 |
| non-user vs regular | 0.37 | 0.33–0.43 | 63% | <0.001 | 0.51 | 0.45–0.58 | 49% | <0.001 |
| irregular vs regular | 0.59 | 0.51–0.67 | 41% | <0.001 | 0.75 | 0.66–0.85 | 25% | <0.001 |
| **Hospitalisation** | | | | | | | | |
| non-user vs user | 0.82 | 0.58–1.14 | 18% | 0.100 | 0.63 | 0.48–0.83 | 37% | 0.001 |
| non-user vs irregular | 1.15 | 0.76–1.69 | -15% | 0.734 | 0.76 | 0.50–1.08 | 24% | 0.143 |
| non-user vs regular | 0.40 | 0.02–1.02 | 60% | 0.038 | 0.00 | 0.00–0.00 | 100% | <0.001 |
| irregular vs regular | 0.35 | 0.02–0.94 | 65% | 0.015 | 0.00 | 0.00–0.00 | 100% | 0.003 |
| **Death** | | | | | | | | |
| non-user vs user | 0.69 | 0.49–0.96 | 31% | 0.011 | 0.57 | 0.40–0.78 | 43% | <0.001 |
| non-user vs irregular | 1.05 | 0.70–1.52 | -5% | 0.586 | 0.72 | 0.46–1.07 | 28% | 0.149 |
| non-user vs regular | 0.22 | 0.01–0.61 | 78% | 0.005 | 0.27 | 0.00–0.74 | 73% | 0.044 |
| irregular vs regular | 0.21 | 0.01–0.62 | 79% | 0.004 | 0.38 | 0.00–1.09 | 62% | 0.212 |

**Table S11**
i-KC22 model with probabilistic stop on infection. Stop probability: 0.30 (irregular), 0.05 (regular). Statistics for hospitalisations and deaths are limited to infected individuals.

| | Simulation | | | | KB22/KC22 | | | |
|---|---|---|---|---|---|---|---|---|
| Summary statistics | RR | 95% CI | Risk Red. | p.val | RR | 95% CI | Risk Red. | p.val |
| **Infection** | | | | | | | | |
| non-user vs user | 0.49 | 0.47–0.51 | 51% | <0.001 | 0.56 | 0.53–0.58 | 44% | <0.001 |
| non-user vs irregular | 1.00 | 0.95–1.05 | 0% | 0.450 | 0.68 | 0.64–0.72 | 32% | <0.001 |
| non-user vs regular | 0.10 | 0.07–0.12 | 90% | <0.001 | 0.51 | 0.45–0.57 | 49% | <0.001 |
| irregular vs regular | 0.10 | 0.07–0.12 | 90% | <0.001 | 0.75 | 0.66–0.85 | 25% | <0.001 |
| **Hospitalisation** | | | | | | | | |
| non-user vs user | 0.82 | 0.58–1.14 | 18% | 0.100 | 0.63 | 0.47–0.84 | 37% | 0.001 |
| non-user vs irregular | 0.76 | 0.51–1.11 | 24% | 0.067 | 0.76 | 0.50–1.05 | 24% | 0.143 |
| non-user vs regular | 1.48 | 0.07–3.84 | -48% | 0.579 | 0.00 | 0.00–0.00 | 100% | <0.001 |
| irregular vs regular | 1.97 | 0.08–5.30 | -97% | 0.768 | 0.00 | 0.00–0.00 | 100% | 0.003 |
| **Death** | | | | | | | | |
| non-user vs user | 0.69 | 0.49–0.96 | 31% | 0.011 | 0.57 | 0.41–0.78 | 43% | <0.001 |
| non-user vs irregular | 0.70 | 0.47–1.01 | 30% | 0.023 | 0.72 | 0.45–1.07 | 28% | 0.149 |
| non-user vs regular | 0.79 | 0.02–2.24 | 21% | 0.301 | 0.27 | 0.00–0.71 | 73% | 0.044 |
| irregular vs regular | 1.15 | 0.02–3.37 | -15% | 0.539 | 0.38 | 0.00–1.05 | 62% | 0.212 |

**Table S12**

i-KC22 model with uniform stop on infection. Stop probability: 1.0 (all individuals). Statistics for hospitalisations and deaths are limited to infected individuals.

### Supplemental Figures

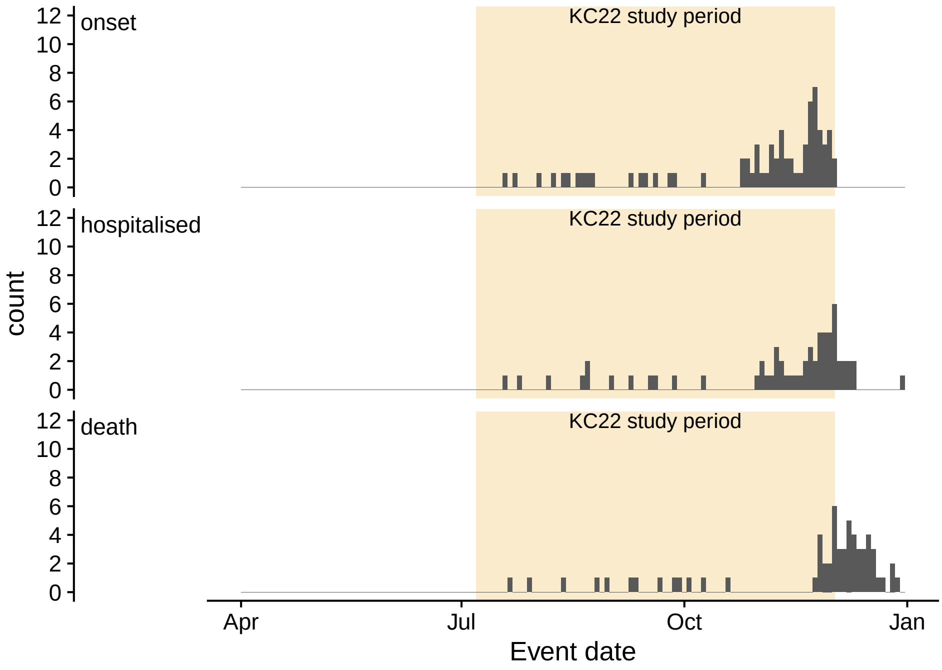


**Figure S1**
**"Missed" deaths.** Event dates for missed Covid deaths after symptom onset during the study period based on matching the SUS data from KC22. The three sub-panels show recorded dates of symptom onset, hospitalisation, and death.

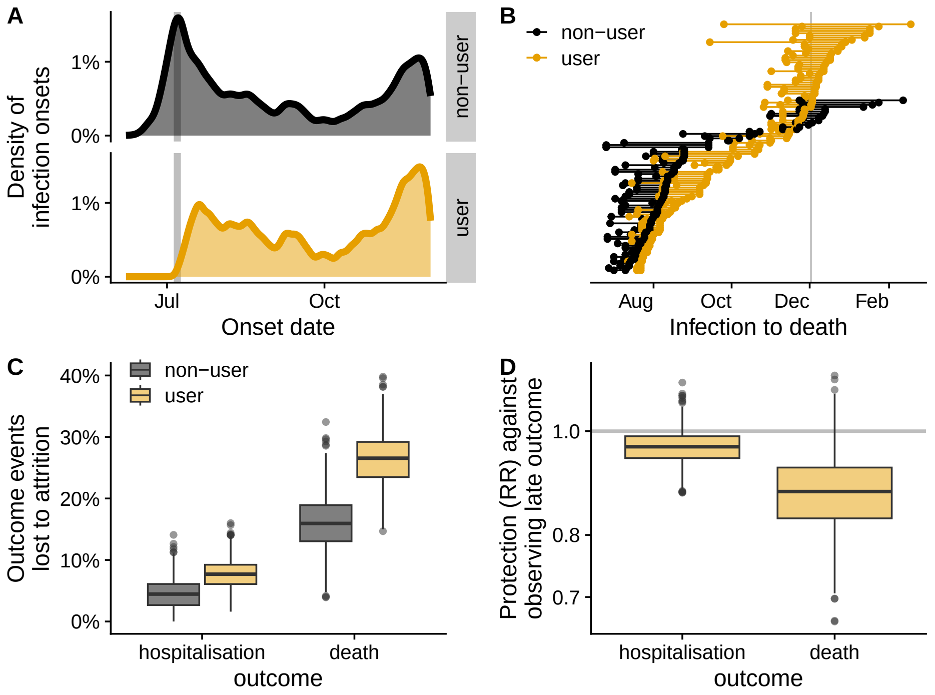


**Figure S2**

**Attrition bias in the i-INF model**. A. Empirical distributions of simulated infection dates over 1000 runs of the i-INF model. Note the delayed early peak of infections in the ivermectin user group. B. Example from one typical simulation of uncounted deaths among ivermectin users. Each line segment represents an individual in the simulation who was infected and later died, with infection and death dates at the end points. The study end date is marked with a vertical line. C. Hospitalisations and deaths are lost to attrition more frequently in the user group (1000 simulations). D. The ivermectin user group has apparent (but fictitious) protection from hospitalisation and death compared to non-users.

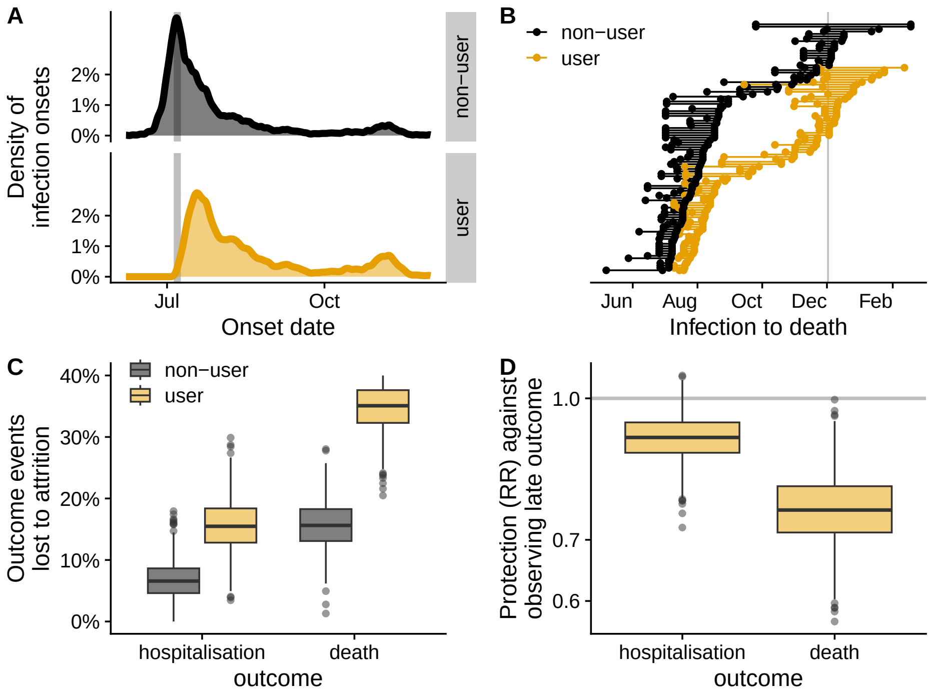


**Figure S3**

**Attrition bias in the i-KC22 model**. A. Empirical distributions of simulated infection dates over 1000 runs of the modified i-KC22 model including late events. Note the delayed early peak of infections in the ivermectin user group. B. Example from one typical simulation of uncounted deaths among ivermectin users. Each line segment represents an individual in the simulation who was infected and later died, with infection and death dates at the end points. The study end date is marked with a vertical line. C. Hospitalisations and deaths are lost to attrition more frequently in the user group (1000 simulations). D. The ivermectin user group has apparent (but fictitious) protection from hospitalisation and death compared to non-users.

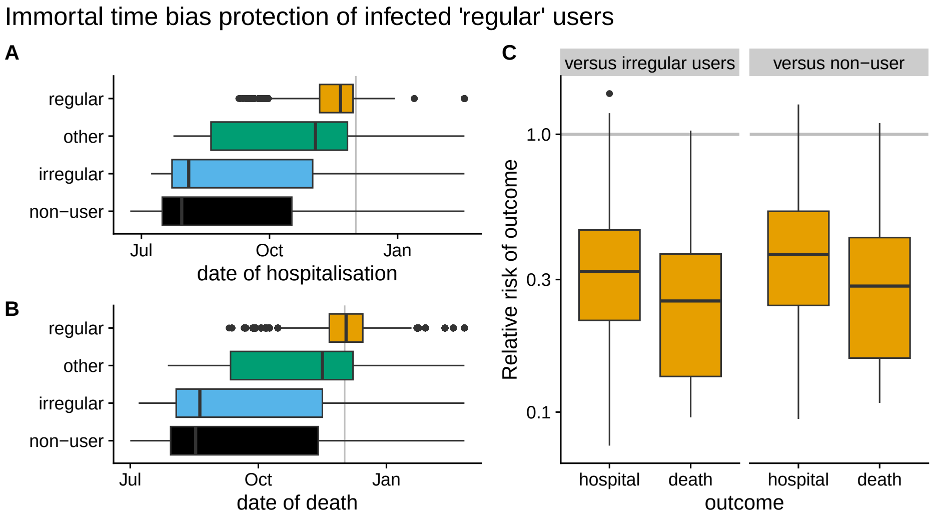


**Figure S4**

**Apparent protection of "regular" users in the i-INF model**. A. Simulated dates of hospitalisation in i-INF for individuals grouped by exposure according to KB22-defined usage groups. The study end date is marked with a vertical line. B. Dates of death. C. Relative risk of hospitalisation and death for "regular" ivermectin users compared to "irregular" and non-users. Risk ratios are calculated and plotted for each of 1000 simulations. Dates of hospitalisation and death are shown for all individuals across 1000 simulations.

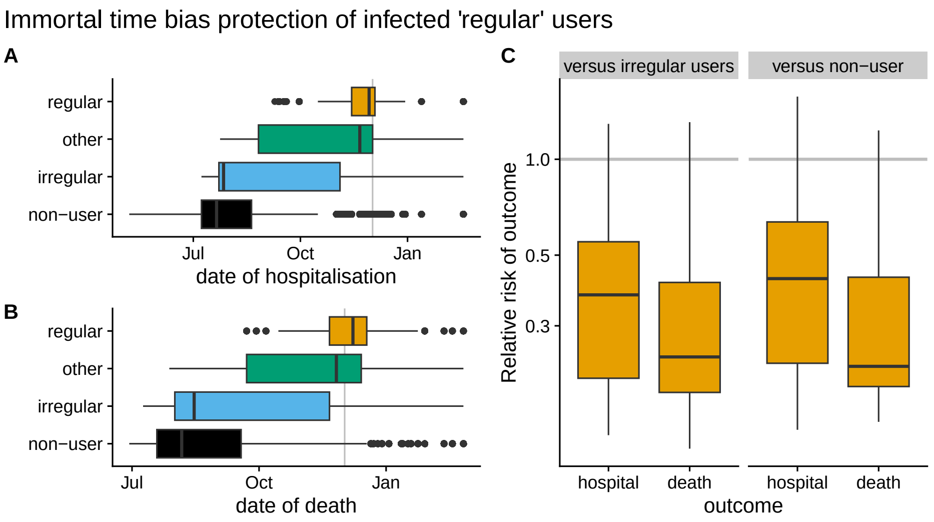


**Figure S5**

**Apparent protection of "regular" users in the i-KC22 model**. A. Simulated dates of hospitalisation in i-INF for individuals grouped by exposure according to KB22-defined usage groups. The study end date is marked with a vertical line. B. Dates of death. C. Relative risk of hospitalisation and death for "regular" ivermectin users compared to "irregular" and non-users. Risk ratios are calculated and plotted for each of 1000 simulations. Dates of hospitalisation and death are shown for all individuals across 1000 simulations.
